# Biallelic variants in *RNU2-2* cause a remarkably frequent developmental epileptic encephalopathy

**DOI:** 10.1101/2025.09.02.25334957

**Authors:** Adam Jackson, Alexander JM Blakes, Elizabeth Wall, Natasha Clarke, Ola Abdelhadi, Shakti Agrawal, Ed Blair, Angela F. Brady, Helen Brittain, Kate E. Chandler, Nicholas Drinkall, Frances Elmslie, Lisa Ewans, Andrew Fennell, Gabriella Gazdagh, Usha Kini, Rebecca Macintosh, Sahar Mansour, Lara Menzies, Kay Metcalfe, Alison Milhench, Elizabeth Palmer, Amitav Parida, Katrina Prescott, Melody Redman, Alessandra Renieri, Rocio Rius, Caterina Lo Rizzo, Rani Sachdev, Cas Simons, Sanjay Sisodiya, Helen Stewart, Huw Thomas, Flavia Tinella, Suzi Walker, Nicola Whiffin, Raymond T. O’Keefe, Jenny Lord, Siddharth Banka

## Abstract

Neurodevelopmental disorders (NDDs) affect 2-4% of the population, are predominantly genetic, and remain unsolved in ∼50% of individuals. We show that rare biallelic variants in *RNU2-2* are enriched and over-transmitted in individuals with unresolved NDDs. We define a novel recessive *RNU2-2* syndrome, delineate its unique genetic architecture and show that clinically it manifests as a severe developmental epileptic encephalopathy. We find that candidate biallelic variants are significantly correlated with reduced U2-2 abundance, implicating compromised transcript stability as likely pathomechanism. We identify decreased ratio of U2-2 to its paralog U2-1 as a potential diagnostic biomarker for this condition. We show that the recessive *RNU2-2* syndrome is genetically, clinically, and mechanistically distinct from the dominant *RNU2-2* disorder. Within our cohort, the recessive *RNU2-2* syndrome emerges as by far the most frequent recessive NDD, greatly disproportionate to the small genomic footprint of this non-protein coding gene.

## INTRODUCTION

Neurodevelopmental disorders (NDDs) are a group of aetiologically and phenotypically heterogenous conditions which affect brain development, leading to early-onset neurological, cognitive and behavioural phenotypes with lifelong impact^1^. NDDs are estimated to affect 2-4% of the population^2^. Developmental and epileptic encephalopathies (DEEs) are a sub-group of severe NDDs characterised by frequent seizures and epileptiform activity, developmental delay or regression, and comorbidities including movement, musculoskeletal, gastrointestinal and sleep disorders^3^. The aetiology of NDDs and DEEs is frequently genetic, with accurate genetic diagnosis underpinning their clinical management from treatment to prognosis, genetic counselling and reproductive decision making^4^. However, even with genome sequencing, almost half of NDD and DEE cases remain unsolved^5,6^.

The major and minor spliceosomes are dynamic complexes of proteins and small nuclear RNAs (snRNAs)^7^ which catalyse splicing in the majority of intron-containing genes. Five vertebrate snRNAs (U1, U2, U4, U5, and U6) are incorporated into the major spliceosome, and all but U5 have paralogs with equivalent roles in the minor spliceosome (U11, U12, U4atac, and U6atac respectively). U5 is incorporated into both the major and minor spliceosomes^8^. Variants in several of the snRNA genes are known to cause human genetic conditions, including NDDs (**Supplementary Table 1**). Recently, we^9^ and others^10^ discovered a relatively frequent dominant NDD caused by heterozygous variants in *RNU2-2*.

*RNU2-2* is a single exon gene located on human chromosome 11. It encodes the 191nt U2-2 snRNA, one of two functional U2 paralogs in humans. U2 recognises the splicing branch site through RNA-RNA base pairing during spliceosomal assembly and facilitates nucleophilic attack of the branchpoint adenosine to the 5’ splice site, in the first step of intron lariat formation^11^. U2-1, a paralog of U2-2, differs from U2-2 at 8 nucleotide positions and is encoded in a 6.1kb tandem repeat array of 5 to 82 identical copies of *RNU2-1* on chromosome 17^12^. Until recently *RNU2-2* was annotated as a pseudogene *(RNU2-2P*), but we have shown U2-2 to be more highly expressed than U2-1 in both the developing brain and retina^9^.

Here, we describe a highly prevalent NDD and DEE caused by biallelic variants in *RNU2-2*. We show that rare biallelic variants in *RNU2-2* are enriched and over-transmitted in a cohort of individuals with unsolved NDDs. We compare genome sequencing data from these individuals with biobank-scale population datasets to prioritise candidate disease-causing variants and reveal the distinctive genetic architecture of the disorder. We combine statistical approaches and detailed clinical phenotyping to provide compelling evidence that these variants cause a novel recessive DEE. We leverage transcriptomic data from affected individuals to show that candidate variants are associated with U2-2 transcript depletion and a decreased U2-2:U2-1 transcript ratio, suggesting a loss-of-function pathomechanism. We also show that the dominant and recessive *RNU2-2* disorders are genetically, clinically and mechanistically distinct. Finally, we demonstrate that the recessive *RNU2-2*-related disorder is the most common recessive NDD in our cohort.

## RESULTS

### Rare biallelic variants in *RNU2-2* are enriched and over-transmitted in individuals with unsolved NDD

We hypothesised that snRNAs are attractive candidates for discoveries of novel recessive ‘RNU-opathies’. We, therefore conducted an enrichment analysis for biallelic variants in snRNA genes using aggregated short-read genome sequencing data with statistical phasing from 78,051 individuals in the 100,000 Genomes Project (100kGP)^13^. We quantified the frequency of rare (internal minor allele frequency <0.001) homozygous and/or compound heterozygous variants for all 1,901 snRNAs annotated in GENCODE v32. In total, we identified 1,897 homozygous and 1,692 compound heterozygous variants in 2,486 individuals. We compared the frequency of rare biallelic variants in individuals with unsolved NDD (*N*=6,762) versus all other individuals in this cohort (henceforth referred to as ‘100kGP controls’, *N*=71,289). The 100kGP controls included unaffected relatives, individuals with non-NDD phenotypes, and individuals with previously solved NDD. We identified nominal enrichment for biallelic variants in individuals with unsolved NDD in two out of three snRNAs genes known to be related with recessive disorders including *RNU4-2*^14,15^ (OR 5.75, 95%CI 1.75-17.0, two-sided Fisher’s exact P=0.00225) and *RNU12*^16,17^ (OR 3.84, 95%CI 1.48-8.95, P=0.00325), but not *RNU4ATAC*^18^ (OR 1.56, 95%CI 0.642-3.31, P=0.252), thus validating our approach (**Figure 1a, Supplementary Table 2)**. Strikingly, we also identified significant enrichment for rare biallelic genotypes in *RNU2-2* (NR_199791.1) (all variants: OR 4.40, 95%CI 2.81-56.72, two-sided Fisher’s exact P=4.4×10^−10^; homozygous variants: OR 9.94, 95%CI 4.70-20.93, P=2.74×10^−09^; compound heterozygous variants only: OR 2.82, 95%CI 1.51-4.95, P=7×10^−4^).

**Figure 1.**
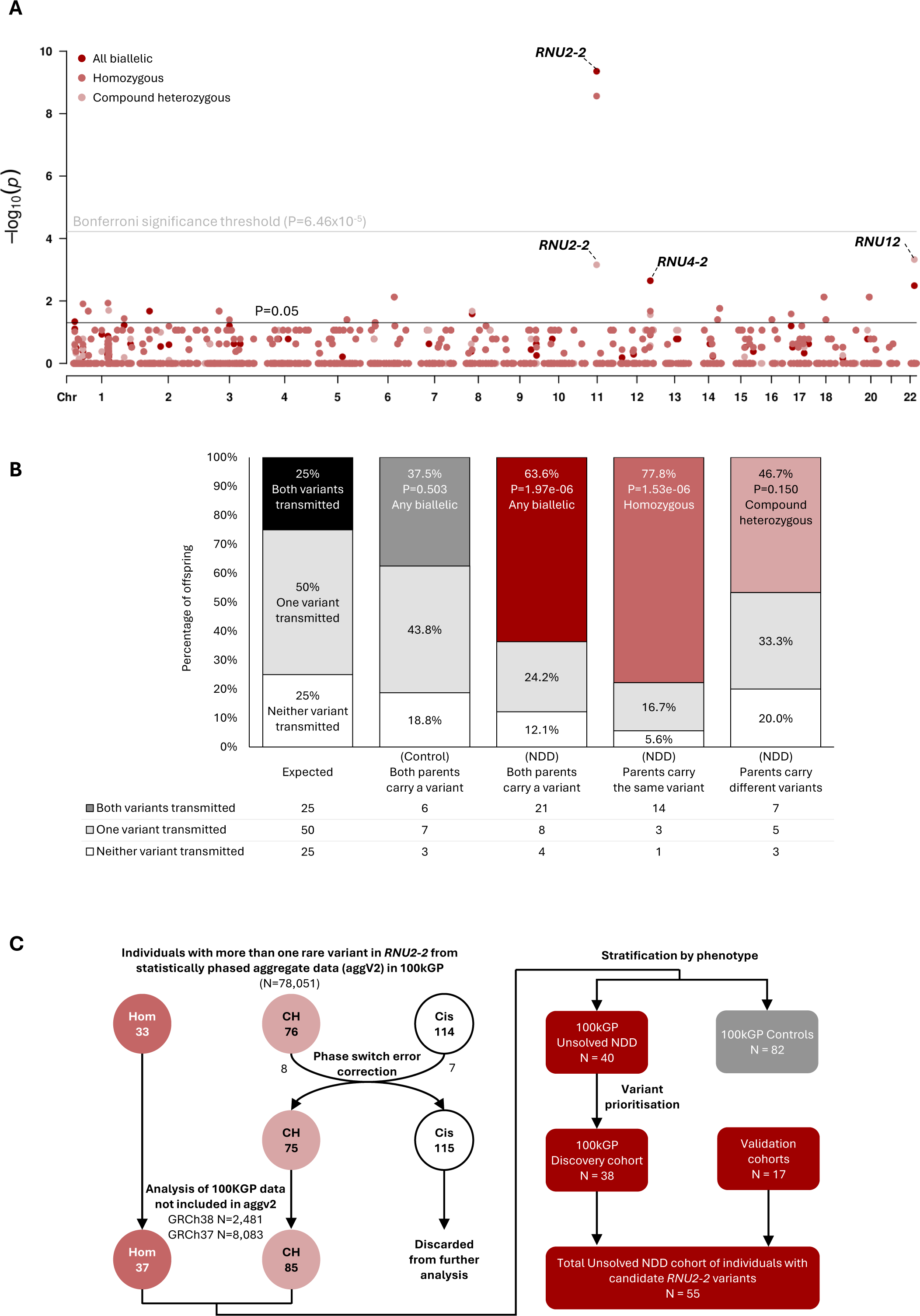
Biallelic variants in *RNU2-2* are enriched and over-transmitted in individuals with unsolved NDD. **A)** Manhattan plot showing biallelic variant enrichment for all snRNAs (*N*=1,901). P values from two-sided Fisher’s exact tests are shown for homozygous variants (light red circles), compound heterozygous variants (pink circles), and all biallelic variants (dark red circles). The Bonferroni significance threshold (P=6.46×10^−5^ for 774 tests at alpha=0.05) is indicated with a grey line. **B)** Stacked bar plot showing transmission of heterozygous variants in *RNU2-2* from parents to offspring. Only trios in which both parents are carriers of heterozygous variants in *RNU2-2* are shown (*N*=49). **C)** Flowchart describing the variant identification and filtering strategy. Hom = homozygous, CH = compound heterozygous, Cis = variants in cis.

In a rare conditions cohort subject to recruitment bias, variants pathogenic for a recessive condition may be over-transmitted to affected offspring^19^. Using genome sequencing data from 12,015 trios in the 100kGP, we identified 49 trios where both father and mother were heterozygous for rare variants in *RNU2-2*. 33/49 offspring from these trios were probands with unsolved NDD (18 homozygotes and 15 compound heterozygotes). We detected a significant over-transmission of *RNU2-2* variants from parents to probands with unsolved NDD versus the expected Mendelian ratios (Chi squared goodness-of-fit P=1.97×10^−6^) (**Figure 1b**). For the remaining 16 trios, where the proband did not have an unsolved NDD, there was no significant deviation from the expected Mendelian ratio for any combination of variants (**Extended Data Figure 1**).

Collectively, the results suggest *RNU2-2* is a likely recessive NDD gene.

### Read-based phasing identifies biallelic variants in *RNU2-2*

From the statistically phased sequencing data described above, we identified 223 individuals with two or more rare variants in *RNU2-2* (33 homozygous, 76 compound heterozygous and 114 with variants *in cis*). Previously, these data were reported to have a 0.18% phase switch error rate for compound heterozygous variants^13^, wherein variants are assigned to the wrong haplotype (**Figure 1c**)^20^. To ensure accuracy of our subsequent analyses, we examined phase switch errors in *RNU2-2* through manual inspection of read alignments for all individuals with more than one rare heterozygous variant in RNU2-2. We identified 15 phase switch errors in total, including seven individuals with variants *in trans* incorrectly phased *in cis*, and eight individuals with variants *in cis* incorrectly phased *in trans*. After accounting for these errors, we identified 33 homozygous and 75 confident compound heterozygous genotypes in 100kGP (**Figure 1c, Extended Data Figure 2**).

The above analysis included only individuals within the statistically phased aggregated variant dataset (aggV2). We searched for additional individuals who were not included in aggV2, totalling 10,564 additional samples: 2,481 aligned to GRCh38 and 8,083 aligned to GRCh37. Because the *RNU2-1* repeat array is not annotated in GRCh37^21^, reads originating from *RNU2-1* are often mapped as low quality *RNU2-2* reads. This reduces the alternate allele fraction of true variants in *RNU2-2* and increases the rate of false negative variant calls. We therefore mapped *RNU2-2* reads from these samples to GRCh38. For individuals with more than one heterozygous variant in *RNU2-2*, we confirmed variant phasing by visual inspection of reads in the Integrative Genomics Viewer (IGV)^22^. Across these additional samples, we identified a further 14 individuals with biallelic variants in *RNU2-2* (four homozygous, ten compound heterozygous; **Figure 1c, Extended Data Figure 3**).

In total, we identified 122 individuals with rare biallelic variants in *RNU2-2* from 100kGP. These included 37 individuals from 30 families with 25 distinct homozygous variants (17 with unsolved NDD and 20 controls) and 85 individuals from 82 families with 80 distinct compound heterozygous genotypes (23 with unsolved NDD and 62 controls) (**Figure 1c, Supplementary Table 3**).

### The distribution of candidate variants is distinct from controls

*RNU2-2* contains four evolutionarily conserved 5’ modules. Stem I forms a four-way helical junction with U6 snRNA and contributes to the stability of the U2-U6 complex^23^. The branchpoint recognition sequence (BPRS) base-pairs with the splicing branchpoint upstream of the 3’ splice site. Stem II undergoes dynamic conformational changes during the splicing process^24^. The Sm binding site is recognised by Sm proteins during snRNP biogenesis^25^. Human *RNU2-2* also possesses two less conserved 3’ modules - Stem III and Stem IV that play a role in 3’ end processing of the pre-snRNA^26^. Visualisation of biallelic variants in the U2-2 primary and secondary structure suggested a clustering of deleterious variants in the 5’ end of the transcript (**Figure 2a, Extended Data Figure 4**a**&b**). Interestingly, we have previously shown that this 5’ region, specifically n.1 to n.60, is constrained for heterozygous variation in population sequencing cohorts^9^.

**Figure 2.**
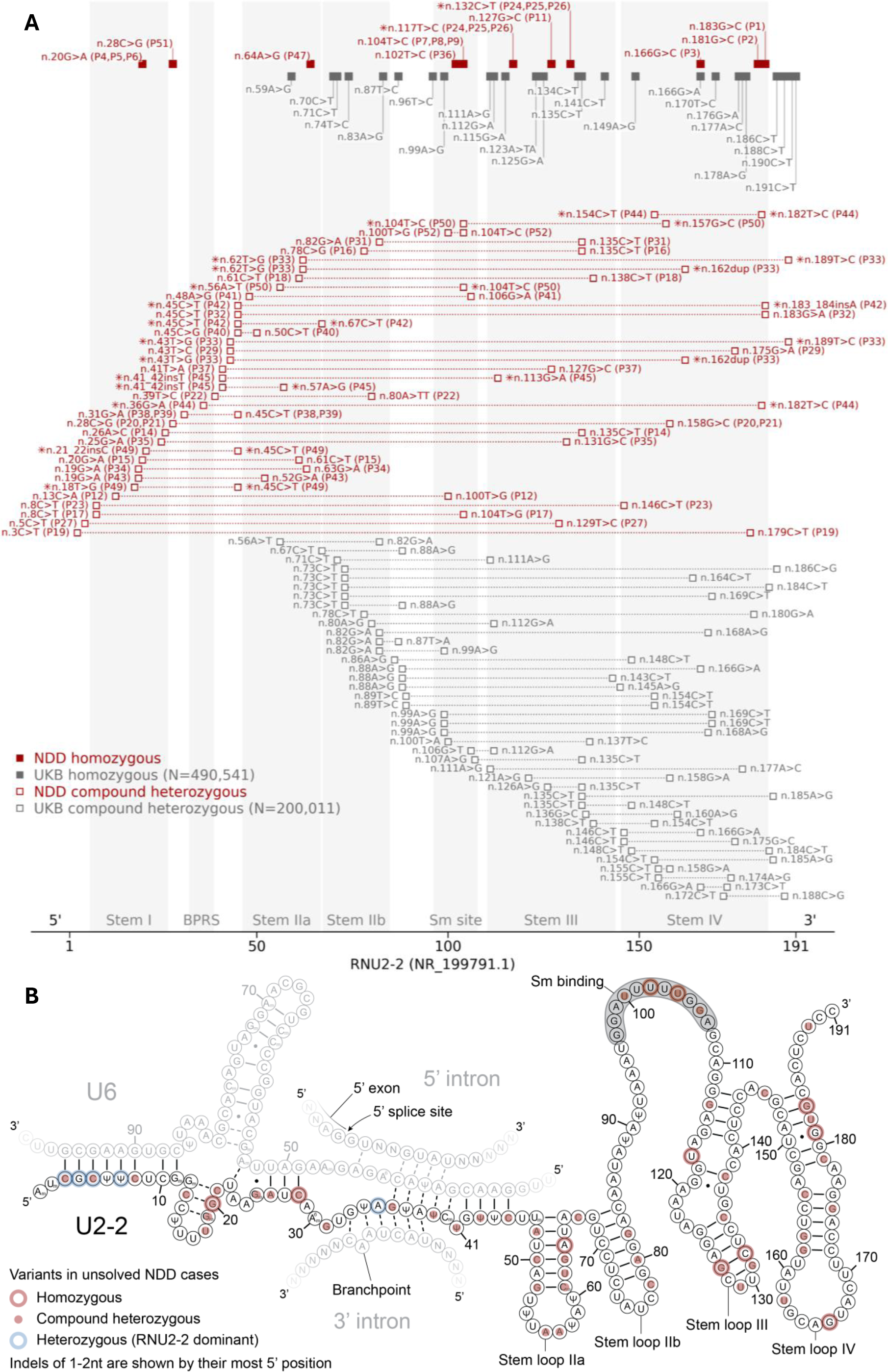
Biallelic variant distributions in *RNU2-2* (NR_199791.1) among unsolved NDD cases and controls. **A)** Transcript diagram showing biallelic variants in *RNU2-2*. Red squares depict candidate biallelic variants in individuals with unsolved NDD from the discovery cohort, GMS, SOLVE-RD and UDNAus. Grey squares show biallelic variants in UKB controls. Closed squares show homozygous variants. Open squares show compound heterozygous variants. For candidate variants depicted in red, the numbers in parentheses show the participant identifier used in this study. Individuals carrying more than two variants are marked with an asterisk. In such cases, all compound heterozygous variant combinations are shown, except for individuals with more than one homozygous variant, for whom only homozygous variants are shown. Only SNVs and indels <3nt in length are shown. Indels are plotted at their most 5’ coordinate. Genotypes which contain variants extending beyond the start or end coordinates of *RNU2-2* are not shown. The extent of the branchpoint recognition sequence (BPRS), Sm site, stem loops I, IIa, IIb, III and IV are highlighted in grey. The boundary of the 5’ constrained region at n.67 is shown by whitespace between stem loop IIa and stem loop IIb. **B)** Secondary structure of U2-2 in complex with U6, the 5’ splice site, and the splicing branchpoint. The proposed four-helix U2/U6 structure described by Karunatilaka and Rueda is shown^51^. Red markers show candidate biallelic variants in unsolved NDD cases. Blue markers show heterozygous variants pathogenic for the dominant *RNU2-2* disorder. The Sm binding site is highlighted in grey. Solid lines show stable Watson-Crick base pairs. Dashed lines show transient Watson-Crick base pairs. Black points show wobble base pairs. 2′-O-methylated nucleotides are depicted by N_m_, pseudouridine by Ψ, and N^6^-methylated adenosine by A^6^. BPRS = branchpoint recognition sequence.

We compared the distributions of biallelic variants in the unsolved NDD cohort versus 100kGP controls and UKB controls within this 5’ constrained region, which we conservatively extend to n.67 to coincide with the junction of stem loop IIa and stem loop IIb (**Figure 2b**). Homozygous variants were scarce in all three cohorts, and we found no significant difference in the number of unique homozygous variants within, or outside of, n.1 to n.67 (NDD vs 100kGP controls OR 1.56, 95%CI 0.0860-28.1, two-tailed Fisher’s exact P=1; NDD vs UKB OR 2.78, 95%CI 0.157-49.2, P=0.484) (**Supplementary Table 4**). However, for compound heterozygous genotypes, we observed a strong enrichment of genotypes with at least one variant within n.1 to n.67 in individuals with unsolved NDD versus both 100kGP controls (OR=34.0, 95%CI 6.81-170, two-tailed Fisher’s exact P=1.07×10^−7^) and 200,011 individuals in UKB for whom statistically phased genome sequencing data was available^13^ (OR 153, 95%CI 19.8-1180, P=1.90×10^−10^). In other recessive RNU-opathies, variants in the Sm binding site are known to be pathogenic^15,27^. However, we did not detect statistical enrichment of homozygous or compound heterozygous genotypes in the Sm site (n.97 to n.107) in individuals with unsolved NDD versus controls (two-tailed Fisher’s exact P>0.0565) (**Supplementary Table 4**).

### Validation across multiple cohorts confirms RNU2-2 as a novel recessive disease gene

To prioritise likely disease-causing variants in our unsolved NDD cohort, we filtered out any individuals with homozygous or compound heterozygous variants that were also observed in population databases (gnomADv4^28^, UK Biobank (UKB)^29^, or All of Us^30^), or in our 100kGP controls in the same combination. Specifically, compound heterozygous genotypes were filtered from the unsolved NDD cohort if the same combination of variants, i.e. the same compound heterozygous genotype, was observed in any control cohort. Ultimately, we prioritised 31 rare biallelic genotypes in *RNU2-2* (10 homozygous and 21 compound heterozygous) in 38 individuals from 31 families with unsolved NDD. These formed our ‘discovery cohort’ in 100kGP (**Figure 1c, Supplementary Table 5**).

To validate our findings, we searched for candidate biallelic variants in *RNU2-2* in additional rare conditions databases. Specifically, we looked for biallelic variants absent in the same combination from UK Biobank, gnomADv4 or 100kGP controls. For compound heterozygous variants, we only included those genotypes in which at least one variant fell in the 5’ constrained region (n.1-n.67) or the Sm site (n.97-n.107). Using these criteria we identified 10 additional individuals in the NHS Genomic Medicine Service (GMS) (*N*=29,872 genomes from 15,889 rare conditions families), two in SOLVE-RD^31^ (*N*=334 genomes) and two in UDNAus^32^ (*N*=249 genomes from 94 families). Notably, although we did not filter by phenotype at this stage, all 14 individuals had unsolved NDD (**Figure 1c**). Finally, we identified three additional affected siblings through segregation analysis.

In total, combining individuals from our discovery and validation cohorts, we identified 55 individuals with NDD phenotypes from 45 families with rare biallelic variants in *RNU2-2* (**Figure 2a, 2b, Supplementary Table 5**), none of whom had another molecular explanation despite prior comprehensive genomic testing.

### The recessive *RNU2-2* syndrome is a developmental epileptic encephalopathy

We examined the phenotypic similarity of 31 unrelated individuals with candidate biallelic variants in *RNU2-2* from our discovery cohort. We excluded seven siblings from this analysis to prevent confounding due to relatedness. For these 31 individuals we compared the Human Phenotype Ontology (HPO) terms documented at recruitment with 1,000 permutations of 31 randomly selected unrelated individuals with NDD in 100kGP. HPO terms in the 31 individuals with candidate variants in RNU2-2 were significantly more homogenous than expected by chance (two-sided Monte Carlo P=0.012, **Figure 3a**).

**Figure 3.**
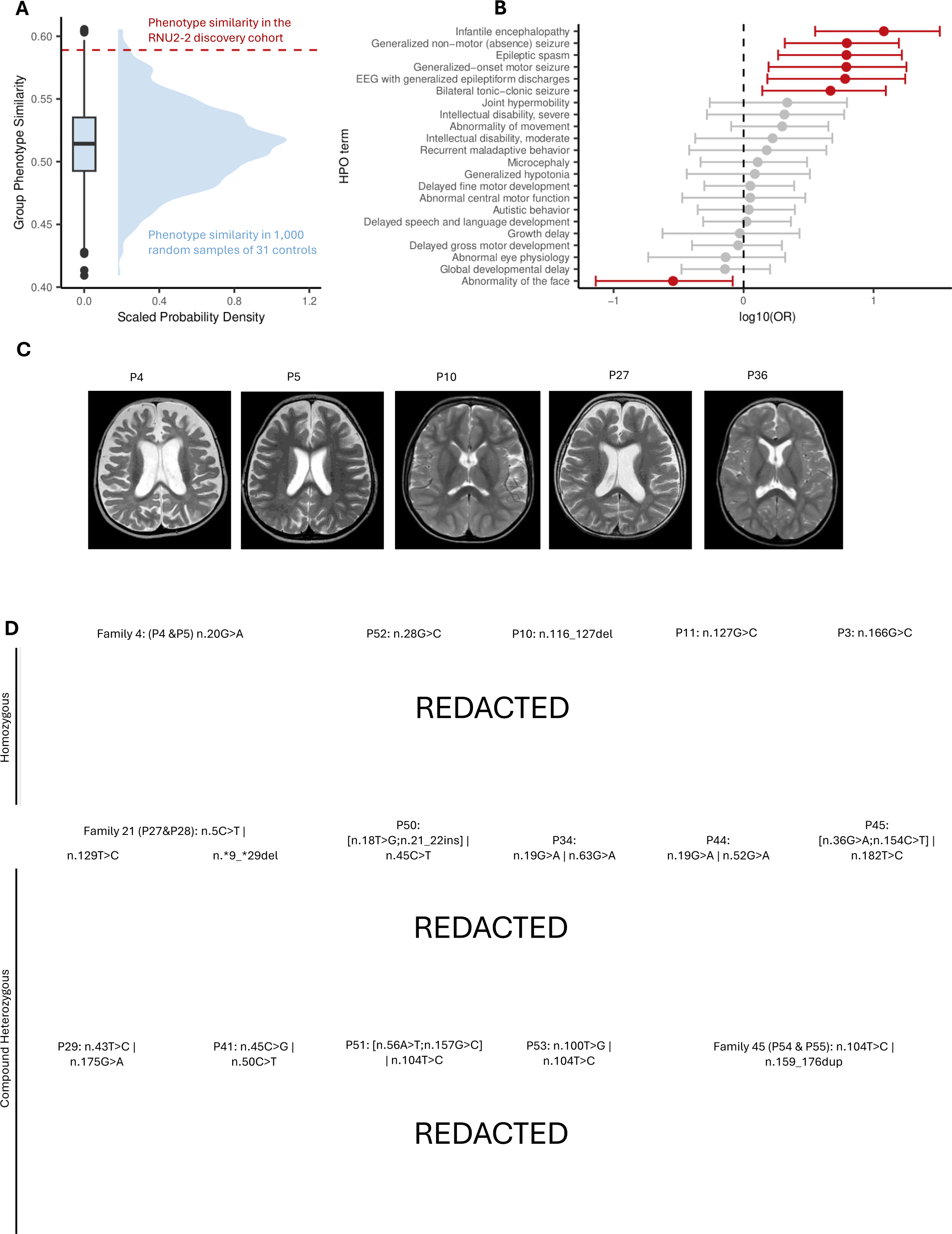
The recessive *RNU2-2* syndrome is a developmental epileptic encephalopathy. **A)** Kernel density estimate and boxplot showing the group phenotypic homogeneity of 1,000 permutations of 31 randomly selected NDD participants in 100kGP versus 31 individuals with candidate biallelic variants in *RNU2-2* (indicated by dashed red line). The boxplot shows the median, quartiles, and +/- 1.5 times the interquartile range of the data. Outliers are shown by filled black circles. **B)** Relative frequency of HPO terms in unrelated individuals with rare biallelic variants in *RNU2-2* versus unrelated 100kGP participants with NDD. HPO terms which differ in frequency between the cohorts are shown in red (two-sided Fisher’s exact tests with Benjamin-Hochberg False Detection Rate (FDR) correction for 71 tests at alpha=0.05, P<0.000704). Error bars show 95% confidence intervals. The raw data are provided in Supplementary Table 7. **C)** T2-weighted axial brain MRI images of individuals with candidate variants in *RNU2-2* showing enlarged CSF spaces and generalised cerebral atrophy (P4, P5, P27 and P36). Prominent cerebral atrophy in the temporal regions is shown for P10. **D)** Facial photographs of individuals with candidate biallelic variants in *RNU2-2*. Participant numbers and variant details in HGVS nomenclature are shown.

To identify the clinical features underlying this phenotypic similarity, we performed HPO term enrichment analysis for these 31 unrelated individuals versus all other unrelated NDD participants in 100kGP (*N*=10,157). Significantly enriched HPO terms were consistent with a DEE, and included “generalised seizures”, “encephalopathy”, “intellectual disability”, and “delayed speech and language development” (**Figure 3b, Supplementary Table 6**).

Next, we performed detailed clinical characterisation of the recessive *RNU2-2* syndrome. We gathered detailed phenotypic information and clinical histories from 28 individuals (21 families) with candidate biallelic variants in *RNU2-2*. 13 were male and 15 were female and their ages ranged between 5 and 34 years (**Supplementary Table 7**, case reports in **Supplementary Note**). All individuals displayed developmental delay and intellectual disability. Most individuals had motor delay (27/28, 96.4%) and seizures (26/28, 92.9%). Seizure onset occurred in the first year of life for 70% (20/26). 79% (22/28) of individuals were non-verbal and 46% (13/28) never developed the ability to walk. Seizure semiology was varied with generalised tonic-clonic seizures in 68% (19/28), myoclonic jerks in 43% (12/28), absence seizures in 29% (8/28) and infantile spasm in 10% (3/28). Encephalopathy was used as a clinical descriptor in 29% (8/28) of individuals. Movement disorders were seen in 35% (10/28) of individuals. Bruxism was noted in 18% (5/28). 57% of individuals required gastrostomy feeding. EEGs were abnormal in 43% (12/28) with epileptiform spike activity in 43% (12/28) and background slowing indicative of encephalopathy in 21% (6/28). MRI scans were abnormal in 46% (13/28) with cerebral atrophy being most common (9/28), followed by enlarged extra-axial CSF spaces in 14% (4/28) (**Figure 3c**). Facial dysmorphism was seen in most individuals (20/28) (**Figure 3d**).

In summary, we demonstrate striking phenotypic convergence in a reverse-phenotyped^33^ cohort with candidate biallelic variants in *RNU2-2* and show that the recessive *RNU2-2* disorder is a developmental epileptic encephalopathy with severe or profound neurodevelopmental delay and prominent speech and gross motor delay.

### Candidate biallelic variants are associated with reduced U2-2 abundance and a decreased U2-2:U2-1 ratio

Recessive inheritance of *RNU2-2*-related DEE suggests loss of function as a likely pathomechanism. Loss of U2 function can occur by several means, including reduced transcript stability^34–38^. To explore the possibility of a loss of function mechanism in our patients, we examined rRNA-depleted RNA sequencing data from blood in 100kGP. We performed expression outlier analysis in 5,412 samples using OUTRIDER^39^. 9/5,412 individuals in this cohort had unsolved NDD and candidate biallelic variants in *RNU2-2* (i.e. belonged to our discovery cohort). 8/9 (89%) of these individuals were significant outliers for *RNU2-2* expression in OUTRIDER (FDR P<0.05) (**Supplementary Table 8**). Only six other individuals in the remainder of the cohort (*N*=5,403) were significant expression outliers for *RNU2-2*. *RNU2-2* Overall, *RNU2-2* expression outliers were highly over-represented among individuals with candidate variants in *RNU2-2* (OR=7,372, 95%CI 795-68,400, P=4.68×10^−15^), suggesting reduced U2-2 expression or reduced transcript stability as a potential mechanism in most individuals with this disorder.

Interestingly, 7/8 individuals with outlier *RNU2-2* expression were also found to be an expression outlier for *WDR74 (***Supplementary Table 8**). *RNU2-2* is located within the 5’ UTR of *WDR74* on chromosome 11 (**Extended Data Figure 5**a), although the two genes are transcribed independently^21^. We reasoned that the overlap of these two loci may confound expression quantification based on read counts. We therefore counted the number of RNA-Seq reads mapping to coding exons of *WDR74* alone and observed no significant difference between cases and controls (mean reads per million (RPM) 96.8 and 85.6 respectively, two-tailed Mann-Whitney U test P=0.151) (**Extended Data Figure 5**b). This implies that the apparent reduced *WDR74* expression in individuals with candidate variants in *RNU2-2* is a technical artefact driven by *RNU2-2* alone.

Next, we explored the transcriptional relationship of U2-1 and U2-2 in cases and controls in these transcriptomic data. First, we counted reads mapping to *RNU2-2* in the nine individuals with candidate biallelic variants, and an expanded set of control samples (*N*=5,443). Consistent with our OUTRIDER results, *RNU2-2* expression was significantly lower for individuals with biallelic variants than controls (mean RPM 102 vs 757, two-tailed Mann-Whitney U test P=1.53×10^−6^) (**Figure 4a, Supplementary Table 9**). Next, we found that expression of U2-1 and U2-2 is highly correlated (Pearson’s R=0.83, P<2.2e-16, **Figure 4b**), consistent with previous results^10^. We did not detect any significant OUTRIDER expression outliers for *RNU2-1* among any individuals in the original cohort of 5,412 samples. Strikingly, all eight *RNU2-2* OUTRIDER outliers among the cases had a significantly depleted ratio of U2-2:U2-1 expression (range 0.352-0.688. No other individuals, including the six *RNU2-2* outliers from the control cohort, had a U2-2:U2-1 ratio in this range (*N*=5,448, range 0.76-1.45). The mean *RNU2-2* ratio was significantly lower for the participants with candidate biallelic variants than controls (0.56 vs 1.04, two-sided Mann-Whitney U test P=4.4×10^−7^) (**Figure 4c**). These data suggest that the U2-2:U2-1 ratio may be a more specific marker for the recessive *RNU2-2* disorder than U2-2 expression alone.

**Figure 4.**
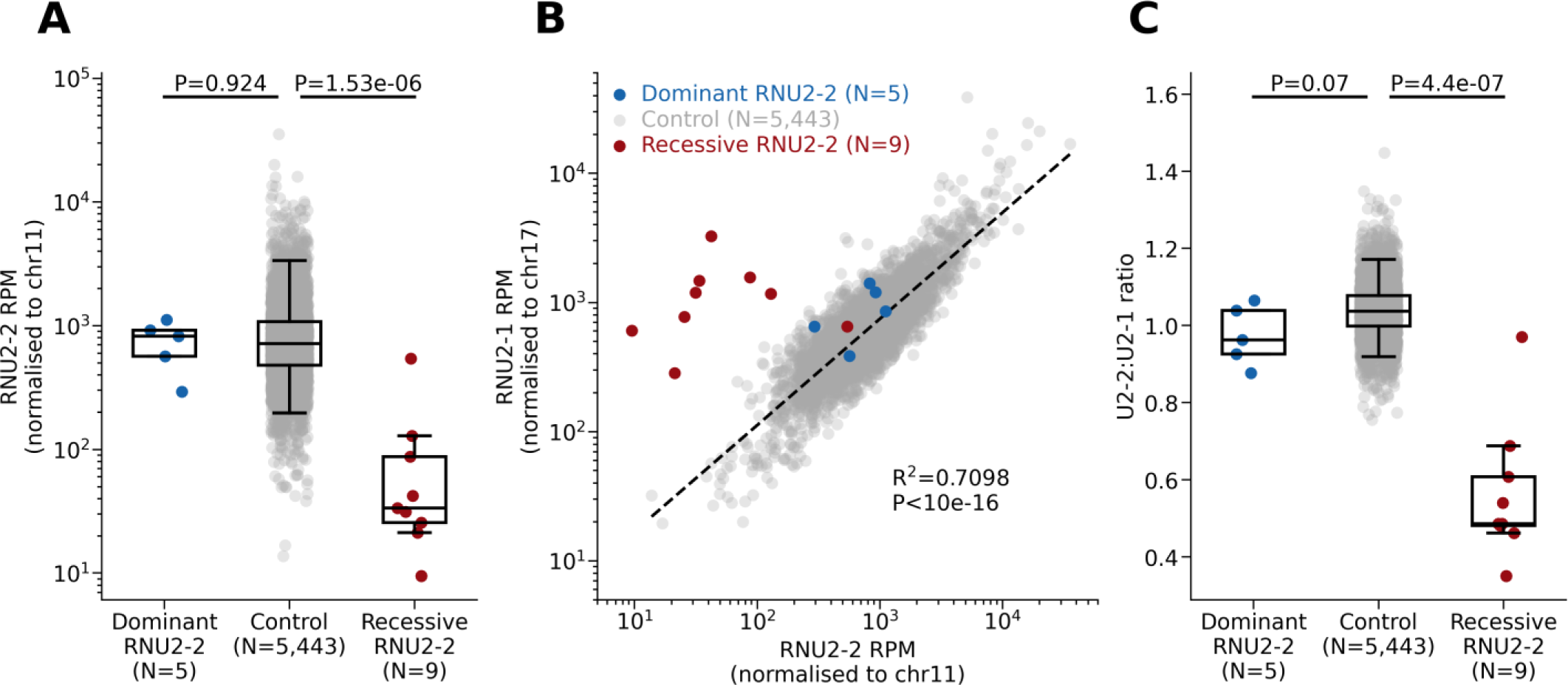
U2-2 and U2-1 transcript abundance in carriers of candidate variants in *RNU2-2* versus controls. **A)** Boxplots showing *RNU2-2* reads per million (RPM), normalised to chromosome 11. **B)** Scatter plot comparing relative expression of *RNU2-2* and *RNU2-1*. The dashed black line shows the ordinary least squares regression line for controls. The R^2^ and P value for this correlation are shown. **C)** Boxplots showing the ratio of U2-2:U2-1 (RPM). Individuals with candidate biallelic variants in *RNU2-2* are shown in red. Individuals with pathogenic heterozygous variants in *RNU2-2* are shown in blue. For **A)** and **C)**, P values for two-tailed Mann-Whitney U tests are shown. Box and whisker plots show the median, quartiles, and +/- 1.5 times the interquartile range of the data.

In summary, these data show that candidate biallelic variants in *RNU2-2* are associated with reduced U2-2 abundance, and specifically a reduced ratio of U2-2:U2-1.

### Dominant and recessive *RNU2-2* syndromes are distinct

We next set out to compare the dominant^9,10^ and recessive *RNU2-2* disorders. The dominant disorder is caused by a limited repertoire of SNVs at the 5’ end of *RNU2-2* (**Figure 2b**), including highly recurrent variants at n.4G>A and n.35A>G. Candidate biallelic variants in the recessive disorder are more widely distributed throughout *RNU2-2*, notwithstanding an enrichment of compound heterozygous alleles at the 5’ end of the gene (**Figure 2a**). Overall, the genotypic spectrum of the recessive disorder appears much broader than the dominant condition.

Next, we compared these disorders at the RNA level, both in terms of the U2-2 transcript itself, as well as its downstream splicing targets. Having shown that candidate biallelic variants are associated with reduced U2-2 abundance and a reduced U2-2:U2-1 ratio, we found that mean U2-2 abundance in individuals with the dominant condition was no different to controls (mean RPM=745 vs 955, two-tailed Mann-Whitney U test P=0.924) (**Figure 4a, Supplementary Table 10**). Similarly, the U2-2:U2-1 ratio in these heterozygous participants did not differ significantly from controls (range 0.88-1.06 vs 0.76-1.08, two-tailed Mann-Whitney U test P=0.07) (**Figure 4c**). These data suggest that U2-2 transcript depletion is likely a specific feature of the recessive disorder.

Whereas deleterious variants in a core component of the major spliceosome may be expected to disrupt splicing, previous work did not identify a splicing defect in blood in the dominant *RNU2-2* disorder^10^. Returning to the transcriptomic dataset described above, we sought to compare the splicing impact of the dominant versus recessive disorders through a splicing outlier analysis with FRASER2^40^. From 5,546 rare conditions participants with transcriptomic data available, we identified five individuals with pathogenic heterozygous variants and nine individuals with candidate biallelic variants in *RNU2-2*. We compared these cases with a subset of 301 controls with non-NDD phenotypes who were under 18 years of age. We did not observe a significant difference in the number of FDR-significant (FDR P<0.10) splicing outliers between groups (**Extended Data Figure 6**a). Following previous work^41^, we reasoned that deleterious variants in *RNU2-2* may have a more subtle impact on splicing. We therefore broadened our analysis to nominally significant (P<0.05) FRASER2 outliers. We found that individuals with the dominant disorder have a significantly greater number of nominally significant splicing outliers in blood than controls (mean splicing outliers 7,072 vs 5,267, two-tailed Mann-Whitney U test P=0.0112), whereas individuals with candidate biallelic variants did not (mean 5,722 vs 5,266, P=0.695) (**Figure 5a**). Because of the prominence of seizures in the clinical presentation of the recessive disorder, we further subset our analysis to nominally significant splicing outliers in known monogenic epilepsy genes (PanelApp Australia v1.178 Genetic Epilepsy panel). Again, we found that individuals with the dominant disorder had a significantly greater number of splicing outliers in these genes than controls (mean 482 vs 350, two-sided Mann Whitney U test P=0.00752), but found no such difference in candidate biallelic cases (mean 358 vs 350, P=0.825) (**Extended Data Figure 6**b).

**Figure 5.**
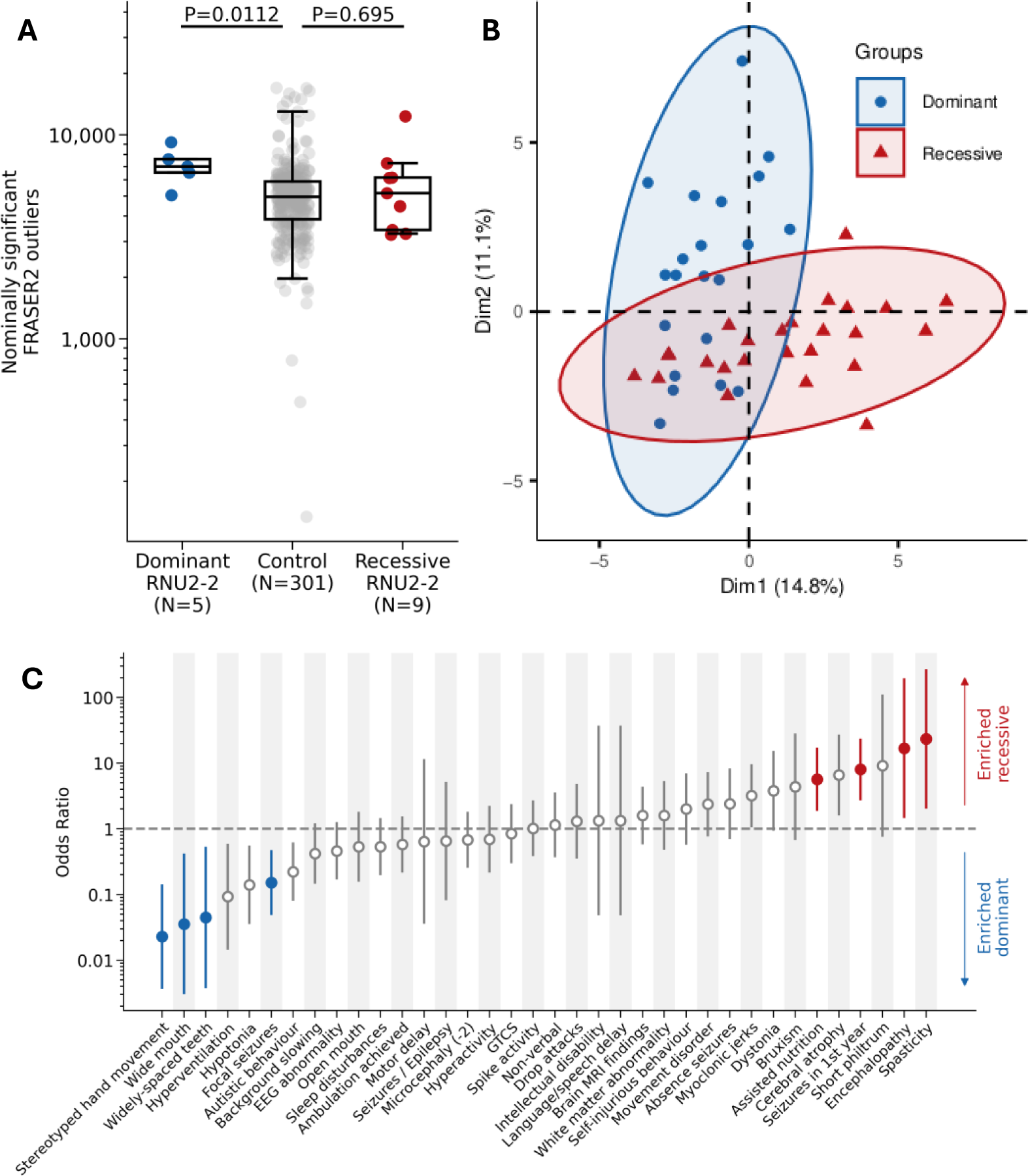
Molecular and phenotypic distinction between the recessive and dominant *RNU2-2* disorders. **A)** Nominally significant FRASER2 outliers (P<0.05) in carriers of candidate variants in *RNU2-2* versus controls. P values from two-tailed Mann-Whitney U tests are shown. Box and whisker plots show the median, quartiles, and +/- 1.5 times the interquartile range of the data. **B)** PCA scatterplot of HPO terms in the recessive and dominant *RNU2-2* disorders. The proportion of the variance explained by each principal component is shown in the axis labels. Ellipses were generated using default parameters of the factoextra R package. **C)** Relative frequency of HPO terms in the recessive and dominant *RNU2-2* disorders. Odds ratios after Haldane-Anscombe correction are shown. Error bars show 90% confidence intervals. Filled circles show FDR-significant P values from two-sided Fisher’s exact tests (alpha=0.05).

We hypothesised that some aberrant splicing events may be shared by individuals with the dominant or recessive disorders. We compared the number of identical, nominally significant aberrant splicing events shared by individuals with candidate variants in *RNU2-2* versus 1000 permutations of an equal number of randomly selected controls. Individuals with the dominant disorder shared a greater number of aberrant splicing events at annotated splice junctions than expected by chance (*N*=1930, two-sided percentile bootstrap P=0.002, Bonferroni P=0.016) (**Extended Data Figure 7**). No such signal was observed for participants with candidate biallelic variants (**Extended Data Figure 8**). In summary, we were able to detect an aberrant splicing signal in blood for individuals with the dominant disorder, but not the recessive disorder.

Finally, we compared the clinical phenotypes of the dominant and recessive conditions using detailed clinical information 26 from individuals with the recessive *RNU2-2* NDD, and published clinical data for 21 individuals with the dominant *RNU2-2* disorder^9,10^. We limited participants with the dominant condition to those carrying the highly recurrent n.4G>A, n.35A>G and n.35A>C variants, because only these variants reach the threshold for a (Likely) Pathogenic classification by current ACMG criteria^42^. Principal component analysis of 46 clinical features showed limited separation of the dominant and recessive *RNU2-2* conditions (**Figure 5b, Extended Data Figure 9**). We therefore compared the frequency of individual phenotype terms between the recessive and dominant disorders. The most strongly enriched clinical features for the recessive disorder included “spasticity” (OR 23.3, 90%CI 21.3-245, two-tailed Fisher’s exact FDR P=0.024), “encephalopathy” (OR 16.8, 90%CI 1.45-195, FDR P=0.038), “seizures in first year” (OR 8.00, 90%CI 2.70-23.7, FDR P=0.017) and “assisted nutrition” (OR 5.67, 90%CI 1.87-17.2, FDR P=0.042; **Figure 5c, Supplementary Table 11**). The most strongly enriched phenotypes for the dominant disorder included “stereotyped hand movements” (OR 0.023, 90%CI 0.00365-0.142, FDR P=3.10×10^−4^), dysmorphic facial features such as “wide mouth” (OR 0.0357, 90%CI 0.00303-0.421, FDR P=0.0173), and “focal seizures” (OR 0.152, 90%CI 0.0483-0.475, FDR P=0.0348; **Figure 5c**).

Taken together, these data suggest that the dominant and recessive *RNU2-2* disorders are genetically, molecularly, and clinically distinct.

### Biallelic variants in *RNU2-2* cause the most common recessive NDD in 100kGP

Finally, we estimated the frequency of the recessive *RNU2-2* disorder compared to other recessive NDDs. We compared the number of individuals with candidate biallelic variants in *RNU2-2* versus the number of individuals with a confirmed recessive NDD using Exit Questionnaire data from 10,157 individuals with NDD in 100kGP. The recessive *RNU2-2* disorder is by far the most frequent recessive condition in this cohort (*N*=31), observed in over three times as many individuals as *VPS13B* (Cohen syndrome, MIM #216550, *N*=9), the next most frequent diagnosis (**Figure 6a**). Expanding our analysis to all NDD genes with “green” review status in the Intellectual Disability panel (R29) in PanelApp^43^, we find that the recessive *RNU2-2* disorder is the second most frequent diagnosis after *RNU4-2*-related ReNU syndrome (*N*=59), and the only recessive disorder among the top 20 most frequent diagnoses in this cohort (**Extended Data Figure 10**).

**Figure 6.**
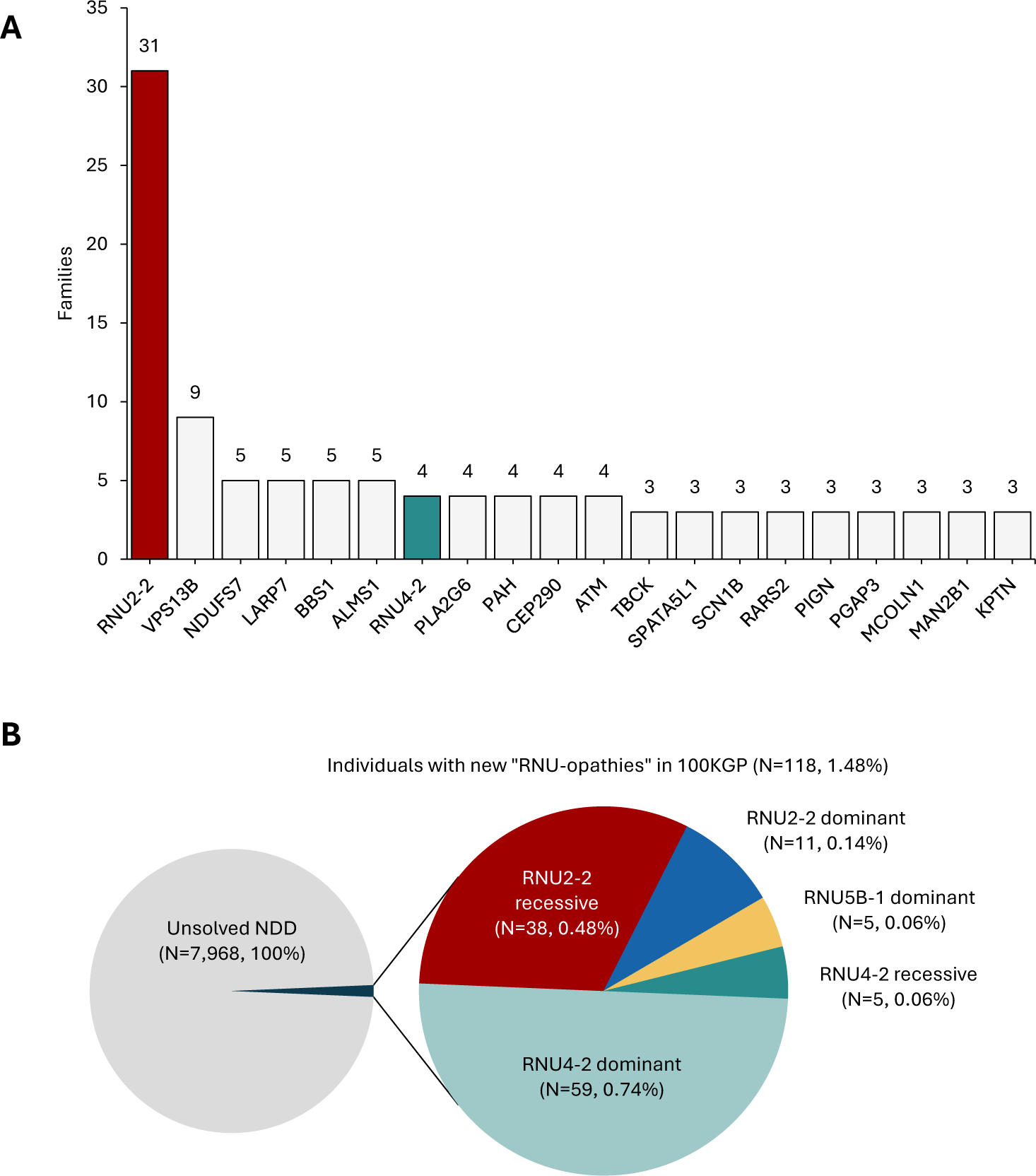
The frequency of the recessive *RNU2-2* syndrome and other “RNU-opathies” in 100kGP. **A)** Bar plot showing the 20 most frequent recessive diagnoses in the rare disease arm of 100kGP. The number of families “solved” by biallelic variants in each gene is shown. Counts for the *RNU2-2* and *RNU4-2* disorders were based on genotyping (see Methods). Counts for all other genes were taken from exit questionnaire data. **B)** Pie chart showing the number of individuals with unsolved NDD in 100kGP explained by pathogenic variants in the recently described “RNU-opathy” genes. Note that panel **A)** shows counts of families, whereas panel **B)** shows counts of individuals.

We previously identified 11 individuals with the dominant *RNU2-2* syndrome in 100kGP^9^. Now, the discovery of 38 individuals with the recessive *RNU2-2* syndrome brings the total number of individuals in this cohort with an *RNU2-2*-related diagnosis to 49/7,968 (0.61%).

The recently discovered RNU-opathies (dominant and recessive *RNU4-2*^14,15,41,44^, dominant and recessive *RNU2-2*^9,10^, and dominant *RNU5B-1* syndromes^9^) account for 118/7,968 (1.48%) individuals with previously unsolved NDDs in 100kGP (**Figure 6b**). This is particularly remarkable as these genes have a combined genomic footprint of 448 nucleotides (i.e. 1 in 1.35×10^7^nt of GRCh38).

In summary, biallelic variants in *RNU2-2* are a highly frequent unrecognised cause of NDD with seizures.

## DISCUSSION

Here, we describe a novel NDD caused by rare biallelic variants in *RNU2-2*, define the genetic architecture of the disorder, characterise its clinical presentation, show that most candidate variants likely result in transcript depletion, identify an RNA-based potential diagnostic biomarker for the condition and find that the recessive *RNU2-2*-related syndrome is the most common recessive NDD in 100kGP.

*RNU2-2* is a single exon non-coding gene of only 191nt. Given this, it is remarkable that variants in this gene are responsible for such a frequent recessive disorder. There is also considerable allelic heterogeneity amongst affected individuals, which is unexpected for such a small gene. Among our candidate cases in 100kGP, compound heterozygous genotypes were more frequent than homozygous genotypes and we did not identify a high number of consanguineous families. The frequency and distribution of compound heterozygous variants among our cases suggests that specific variant combinations are important to the aetiology of the disorder. We hypothesise that the recessive *RNU2-2*-related syndrome occurs within a specific window of residual U2-2 function, such that a combination of two strongly deleterious alleles may be non-viable, while two mildly deleterious alleles may not cause a clinically obvious phenotype. In this scenario, alleles which are pathogenic in the homozygous state may have intermediate deleteriousness. We note that homozygous variants are very scarce in the 5’ end of *RNU2-2*, whereas this region is enriched for compound heterozygous variants in individuals with NDD. Such a “Goldilocks” phenomenon has precedent in other small non-coding RNA genes associated with recessive conditions, including *RNU7-1* (Aicardi-Goutieres syndrome, #619487)^45^ and *SNORD118* (cerebral microangiopathy and leukoencephalopathy, #614561)^27^. We suggest that this phenomenon may be relevant for other rare conditions and may have been under-recognised thus far.

As in some other snRNA genes^15^, *RNU2-2* has a remarkably high density of rare variants, likely driven by a high mutation rate balanced by negative selection^15,41^. The broad genotypic spectrum of this recessive disorder presents an obstacle to variant interpretation in the clinical setting. Whereas we provide strong statistical and clinical evidence for *RNU2-2* as a novel recessive disorder gene, many of our candidate variants will be classed as variants of uncertain significance according to clinical variant interpretation guidance. The interpretation of non-coding variants is known to be especially challenging^46^. Future studies which leverage larger cohorts of affected individuals and variant-specific functional data will facilitate clinical variant interpretation.

We show that the recessive *RNU2-2*-related syndrome is a DEE characterised by severe to profound global developmental delay, intellectual disability, early onset seizures of varying semiology, movement disorders, requirement of gastrostomy feeding, abnormal electroencephalograms, and evidence of cerebral atrophy on neuroimaging. More detailed natural history studies will be essential to investigate the range and frequency of clinical features and comorbidities caused by this disorder.

In contrast to the dominant disorder, and consistent with a recessive mode of inheritance, we show that candidate biallelic variants in *RNU2-2* are associated with U2-2 transcript depletion, indicative of a loss-of-function mechanism. Similar results have been described for other recessive RNU-opathies. For example, biallelic variants in the *RNU12*, associated with spinocerebellar ataxia 33 (MIM 620208), have previously been shown to reduce U12 stability and transcript abundance^47^. We hypothesise that the transcript depletion we observe may be secondary to reduced transcript stability. Understanding the molecular basis of this disorder will be critical for the development of targeted therapies.

Importantly, we also find that a reduced U2-2:U2-1 ratio is a specific marker for the recessive disorder. U2-2 and U2-1 are found mutually exclusively in the major spliceosome^21^, although whether U2-2 and U2-1 spliceosomes have identical splicing profiles is unknown. These paralogous genes differ at only eight nucleotides, four of which are weakly conserved positions adjacent to the Sm site^9^. It is therefore plausible that U2-1 may partially compensate for under-expression of U2-2. Indeed, previous studies have shown that these genes can functionally compensate for each other^48^. This may be especially true in blood, where the expression of U2-1 relative to U2-2 is greater than in brain or retinal tissue^9^, and the expression of both genes is highly correlated. A reduced U2-2:U2-1 ratio in blood implies either stable U2-1 expression independent of U2-2 levels, or compensatory over-expression of U2-1 in response to U2-2 depletion. Whether *RNU2-1* is dynamically upregulated is unknown. The genetic architecture of the *RNU2-1* locus may also influence its function in a tissue-specific manner; other macrosatellites, such as *D4Z4*, are subject to tissue-specific regulation through complex epigenetic controls^49^.

Deleterious variants in a core component of the spliceosome may be expected to disrupt splicing. Indeed, heterozygous variants in *RNU4-2* pathogenic for ReNU syndrome have been shown to disrupt 3’ splice site selection^41,50^, and a biallelic variant in *RNU12* has been shown to cause minor intron retention in blood^16^. Here, we observed a measurable increase in aberrant splicing events in blood from individuals with the dominant *RNU2-2* disorder, but we see no such defect in the recessive disorder. Because variants which grossly disrupt splicing are likely to be incompatible with life, any splicing defect in the recessive disorder is likely to be subtle and/or tissue specific. We are unable to comment on the effect on splicing in neural tissue as our analysis is limited to short-read RNA-Seq in whole blood. Further RNA studies are warranted to clarify the splicing defect the recessive *RNU2-2* disorder, using other sequencing approaches, patient-derived cell lines, clinically relevant tissues, or *in vitro* and *in vivo* model systems.

Although our findings will need replication in other populations, we found that *RNU2-2* was the most common recessive NDD in the 100kGP. The discovery of a highly prevalent severe recessive disorder is especially significant because accurate carrier testing can inform reproductive decisions and enable preconception screening and prenatal testing. Several factors may explain why the genetic basis of such a prevalent recessive disorder has remained elusive until now. For example, *RNU2-2* is excluded from most exome capture kits. Furthermore, until recently, *RNU2-2* was annotated as a pseudogene (*RNU2-2P*), while the absence of the *RNU2-1* locus from GRCh37 assembly confounded the accurate detection of variants in *RNU2-2*. Finally, the high density of variants in the gene, the lack of founder variants, and the preponderance of compound heterozygous genotypes in this condition reduce the efficacy of traditional recessive disease gene discovery approaches such as autozygosity mapping. Epidemiological studies to determine the incidence, prevalence and the carrier rates for the recessive *RNU2-2* syndrome will be required in future.

In summary, we describe a highly prevalent NDD and DEE caused by biallelic variants in *RNU2-2*. This discovery will enable diagnosis for thousands of individuals with unsolved NDD worldwide and catalyse future research into the biology, epidemiology, and treatment of this condition.

## ONLINE METHODS

### Enrichment analysis for biallelic variants in snRNA genes

A list of snRNAs was generated by filtering the GENCODE comprehensive annotations (v32) for lines containing the string ‘gene_type “snRNA”’. Statistically phased sequencing data, aligned to GRCh38, for 78,051 individuals from 100kGP (AggV2) was accessed within the Genomics England Research Environment (GERE). Biallelic variants in snRNA genes were identified within these data using bcftools (v1.16)^52^ and custom scripts. Only PASS variants were included. Variants occurring above 0.001 allele frequency in AggV2 were filtered.

Individuals were defined to have an ‘unsolved NDD’ if they fulfilled the following criteria: (1) Individuals with any of the following HPO terms: “Global developmental delay” (HP:0001263), “Profound global developmental delay” (HP:0012736), “Severe global developmental delay” (HP:0011344), “Moderate global developmental delay” (HP:0011343), “Mild global developmental delay” (HP:0011342), “Intellectual disability” (HP:0001249), “Intellectual disability, profound” (HP:0002187), “Intellectual disability, severe” (HP:0010864), “Intellectual disability, moderate” (HP:0002342), “Intellectual disability, mild” (HP:0001256), “Intellectual disability, progressive” (HP:0006887), “Intellectual disability, borderline” (HP:0006889), “Autism” (HP:0000717), “Autistic behavior” (HP:0000729), or “Autism with high cognitive abilities” (HP:0000753) as per Chen et al^41^; or (2) were recruited under disease category ‘Intellectual disability’; and (3) “Case solved” status did not equal ‘yes’ in the Genomic Medicine Exit Questionnaire (gmc_exit_questionnaire table in LabKey); and (4) Absent from the “submitted_diagnostic_discovery” table in LabKey.

10,987 individuals in 100kGP were identified who had NDD as defined above. A total of 7968 individuals remained unsolved in v4 release (preceding the publication of *RNU4-2*, *RNU2-2* and *RNU5B-1* dominant syndromes). 6,762 individuals with unsolved NDD were identified within the AggV2 data. The remaining 71,289 individuals in aggV2 were defined as “100kGP controls”. The number of homozygous and compound heterozygous variants in each snRNA gene was calculated for *N*=*N*=both cohorts. For each gene with at least one variant (*N*=774), odds ratios for the number of variants in each cohort were calculated. Statistical significance was determined with two-sided Fisher’s exact tests with Bonferroni correction at alpha=0.05.

### Transmission analysis of rare heterogeneous *RNU2-2* alleles

100kGP trios, *N*=aligned to GRCh38, were identified from the “denovo_cohort_information” table in LabKey (*N*=12,015 trios). Parent pairs where both father and mother were heterozygous for a single rare variant in *RNU2-2* (minor allele frequency <0.001 in aggV2) were identified using custom scripts. The resulting trios were then stratified according to whether parents were heterozygous for the same variant or for different variants, and whether the offspring was a member of our “unsolved NDD” or “100kGP control” cohorts.

The *RNU2-2* genotype of the offspring in each trio was determined. The number of times that either both, neither, or one allele was transmitted to the offspring was counted. A Chi-Squared goodness-of-fit test was performed for the observed number of transmissions against the expected number according to Mendelian ratios.

### Phase switching error detection

All individuals in aggV2 with two or more rare variants in *RNU2-2* were identified using bcftools v1.16^53^. Any variants that occurred in aggV2 at allele frequency greater than 0.001 were removed. IGV^22^ was used to generate screenshots of the entire *RNU2-2* sequence for all individuals with two or more rare variants. These were inspected manually and were either deemed in *trans* if the variants occurred on mutually exclusive reads (with at least one read traversing both variant positions) or in *cis* if variants occurred on the same reads (with at least one read traversing both variant positions).

### Identification of *RNU2-2* variants in 100kGP cases not included in aggV2

A list of genomes not included in aggV2 was generated using the “rare_diseases_participant_disease”, “aggV2_gvcf_sample_stats”, and “genomes_and_file_paths” tables in LabKey. For samples aligned to GRCh38, all variants in *RNU2-2* with FILTER==”PASS” were retained. Samples were then filtered to those carrying either a homozygous variant or more than one heterozygous variant. For samples carrying more than one heterozygous variant, and where at least one variant occurred in n.1-n.67 or the Sm site, phase was determined by manual inspection of reads in IGV.

For samples aligned to GRCh37, all variants in *RNU2-2* with FILTER==”PASS” were extracted. For samples carrying more than one heterozygous variant, all reads aligning to *RNU2-2* (plus 100bp upstream and downstream) were extracted from BAM files. These reads were converted to FASTQ format using BEDtools (v.2.31.0)^54^ and realigned using Bowtie2 (v.2.5.2)^55^ to the GRCh38 FASTA sequence of *RNU2-2*. Only reads with MQ>30 were kept. Variant phase was determined by manual inspection of reads in IGV.

### Curation of biallelic variants in *RNU2-2* in control databases

Homozygous variants in gnomADv4 and the All of Us dataset were reviewed in respective online browsers (https://gnomad.broadinstitute.org/ and https://databrowser.researchallofus.org/, respectively). Homozygous variants in *RNU2-2* were also identified in short-read genome sequencing data 490,541 individuals in UK Biobank^29^. Compound heterozygous variants were identified from a subset of 200,011 of these individuals for whom statistically phased genotype data was available^56^.

### Variant position enrichment analysis

Biallelic genotypes were identified among our unsolved NDD cohort, 100kGP controls and controls in UKB as described above. The statistical analysis of variant distributions was limited to genotypes in which both alleles were entirely contained within the start and end coordinates of the canonical *RNU2-2* transcript (NR_199791.1), and in which both variants were either SNVs or indels less than 3nt in length. The odds ratio that genotypes included a variant within the 5’ constrained region (n.1 to n.67) or the Sm site (n.97 to n.107) of *RNU2-2* was calculated for cases versus controls. P values were determined from two-sided Fisher’s exact tests.

### Cohort expansion

The Genome Medicine Service (GMS) data was accessed through the genome_file_paths_and_types table in LabKey. Variants from 29,782 samples were extracted and filtered as described above for the 100kGP cohort. Compound heterozygous genotypes were identified as described above.*N*= The SolveRD data (*N*=334 genomes) was accessed through the RD-CONNECT portal (https://rd-connect.eu/). Variants were filtering using the Genotype-Phenotype Analysis Platform (GPAP) with gnomAD MAF <0.001 and for non_coding_exon_variant consequence. Individuals with two or more heterozygous genotypes were retained if at least one variant fell within n.1-n.67 or the Sm site of *RNU2-2*. Compound heterozygosity was inferred by manual inspection of reads in IGV.

The UDNAus dataset (*N*=249 genomes from 94 families) was accessed through the Centre for Population Genomics’ rare disease genomic analysis platform, CaRDinal. Variants were filtered for those with gnomADv4 MAF <0.001 using seq^32^. Variants were phased using parental data and compound heterozygous or homozygous genotypes were retained if at least one variant fell within positions n.1–n.67 or the Sm site of RNU2-2.

### Phenotype similarity analysis and enrichment of HPO terms

HPO terms were extracted for all probands in our discovery cohort. Siblings (*N*=7) were excluded in order to limit this analysis to unrelated individuals only (*N*=31). Phenotype similarity was computed using the OntologySimilarity package^57^. We sampled 1000 permutations of the same number (*N*=31) of randomly selected unrelated individuals with NDD in 100kGP. Kernel density estimate and boxplots were plotted using ggplot2 in R and the Monte Carlo P value was calculated for the observed similarity statistic.

To calculate enrichment of HPO terms in the recessive condition, HPO terms were extracted for the 31 probands in our discovery cohort. The OntologyX^57^ R package was used to determine HPO term frequencies in the cohort and remove redundant terms. This was repeated for all unrelated individuals with NDD in 100kGP (*N*=10,157). Odds ratios for non-redundant terms between the two cohorts were calculated. Significance was determined with two-sided Fisher’s exact tests followed by FDR correction.

### Clinical characterisation of recessive *RNU2-2* disorder

This research was performed under the ethical approvals given by the South Manchester National Health Service (NHS) Research Ethics Committee (REC; 11/H1003/3/AM02). Written informed consent for the inclusion of detailed clinical information, imaging data, and facial photographs, was obtained from all participants or their parents.

Contact was made with recruiting clinicians through the Genomics England Airlock (Contact Clinicians Request). A standardised clinical proforma was circulated to all clinicians for completion.

### RNA sequencing and aberrant splicing analyses

Whole blood samples were collected from a subset of 100kGP participants at the time of recruitment and stored in PaxGene tubes for future RNA-Seq. The protocol for RNA sequencing and data processing is publicly available here: https://re-docs.genomicsengland.co.uk/rna_seq/. Briefly, RNA was extracted from whole blood samples from 5,546 probands with rare conditions. Samples were depleted of ribosomal RNA and sequencing libraries were constructed using the Illumina Stranded Total RNA Prep with Ribo-Zero Plus protocol. Sequencing was performed with 2×100bp paired-end reads on a NovaSeq 6000. Read mapping was performed using the Illumina DRAGEN RNA Pipeline v3.8.4.

Splicing outlier analysis with FRASER2^40^ and gene expression outlier analysis with OUTRIDER^39^ were performed for 5,412 samples in batches of 500 samples as part of the DROP pipeline^58^. Significant outlier events in FRASER2 were defined as those with an FDR-adjusted P value <0.1. Nominally significant outlier events were defined as those with an unadjusted P value <0.05. We identified five individuals with pathogenic heterozygous variants in *RNU2-2*, and nine individuals with candidate biallelic variants in *RNU2-2* within this cohort. FRASER2 splicing outliers within these case sets were compared against 301 controls, defined as participants under 18 years of age with non-NDD phenotypes. Specifically, we defined individuals with non-NDD phenotypes as those not recruited under the “Neurology and Neurodevelopmental Disorders” Disease Group in 100kGP (as per the “rare_disease_participant_disease” table in LabKey), and who did not have any of the following HPO terms: HP:0000729 (autistic behaviour), HP:0001250 (seizure), HP:0000252 (microcephaly), HP:0000750 (delayed speech and language development), and HP:0001263 (global developmental delay). Subsequently, these analyses were repeated for a subset of splicing outliers within known monogenic epilepsy genes with an “amber” or “green” review status on the Genetic Epilepsy panel in PanelApp Australia v1.178.

To identify shared splicing events between *RNU2-2* variant carriers, the number of nominally significant FRASER2 outlier events observed in more than one individual was counted. The number of shared splicing events was then counted for 1,000 permutations of an equal number of randomly selected controls (*N*=5 vs heterozygous cases, *N*=9 vs biallelic cases). This procedure was performed for each class of aberrant splicing event (annotated splice acceptor and splice donor, novel acceptor and annotated donor, novel donor and annotated acceptor, novel acceptor and novel donor). Significance was tested with two-sided percentile bootstrap P values followed by Bonferroni correction for eight tests at alpha=0.05.

### Quantifying *RNU2-2* and *RNU2-1* expression

Expression outlier analysis was initially conducted with OUTRIDER from the DROP pipeline run on 5,412 samples, as described above. Significant outlier events in OUTRIDER were defined as those with an FDR-adjusted P value <0.05.

To directly quantify *RNU2-2* and *RNU2-1* expression, reads aligning uniquely to U2-2 (located on chromosome 11) and U2-1 (including all 13 copies of *RNU2-1* located on chromosome 17 in the GRCh38 reference genome) were counted using Samtools^53^. RPM values for *RNU2-2*, WDR74, and *RNU2-1* were obtained by normalising read counts to the total number of unique reads aligning to chr11 or chr17, respectively. For *RNU2-2* and *RNU2-1*, this analysis was performed on 5,457 samples for which RNA-Seq data were available in BAM format. An identical procedure was performed for *WDR74* at a later timepoint on a slightly larger cohort of 5,544 samples. Two-tailed Mann-Whitney U tests were used to compare RPM values and U2-2:U2-1 ratios between cases and controls.

### Comparison of phenotypes of dominant and recessive U2-related disorders

Detailed clinical information, with written consent in place, was obtained through personal correspondence with the responsible clinician for 28 individuals with candidate biallelic variants in *RNU2-2*. For the dominant disorder, detailed clinical information was obtained from the supplementary materials of Jackson et al^9^ and Green et al^10^.

Phenotype terms were harmonised across all returned proformas. For principal components analysis, phenotypes were encoded as Boolean values as per Nava et al.^50^ and Rius et al.^15^. No distinction was made between different severities of the same phenotype (e.g. global developmental delay). Missing values were coded as absent, i.e. “0”. Principal component analysis was performed in R using the FactoMineR^59^ and factoextra^60^ packages.

The enrichment of phenotype terms between the recessive and dominant disorders were quantified with odds ratios. Odds ratios were only calculated for phenotypes observed in at least five individuals in either the dominant or recessive cohorts. Where phenotype counts included zero values, the Haldane-Anscombe correction was applied. Statistical significance was determined with two-sided Fisher’s exact tests with FDR correction.

### Estimating the frequency of the recessive *RNU2-2* disorder and its comparison with other NDDs

Genomic Medicine Centre Exit Questionnaires were accessed from the gmc_exit_questionnaire table in LabKey within the GERE. NDD cases were defined using the rare_disease_participant_phenotype table in LabKey, retaining all individuals who had phenotype terms matching those used for the enrichment analysis, above. The exit questionnaire data were filtered for these individuals only. The exit questionnaire data were filtered for genes with “green” review status within the Intellectual Disability panel (R29) in PanelApp^43^. The number of families with a confirmed diagnosis attributed to each gene was obtained. The number of *RNU4-2*-related ReNU syndrome diagnoses was determined by counting variants in the 18bp critical region reported by Chen et al^41^ for all individuals with NDD. The number of dominant *RNU2-2* diagnoses was determined by counting the recurrent n.4G>A, n.35A>G, and n.35A>C variants among all individuals with NDD. The number of *RNU4-2* recessive diagnoses was taken from the prioritised 100kGP biallelic variant carriers described in Rius et al^15^.

## ACKNOWLEDGEMENTS

We thank the participants and the recruiting clinicians of the 100kGP. We thank Genomics England for generating the data and providing the GERE platform for analysis. We thank Peter O’Donovan, Mitra Sato and Zainab Mustafa for airlock requests, Mimoza Hoti, Anna Lisa Taylor and Joanne Yang for facilitating clinical collaboration requests, and Meriel McEntagart, Cassandra Smith, and Nour Elkhateeb at Genomics England for additional support. We thank Joachim De Jonghe for his suggestions on the secondary structure illustration.

A.J. and S.B. acknowledge the support of SolveRD. The SolveRD project has received funding from the European Union’s Horizon 2020 research and innovation program under grant agreement 779257. This study has been delivered through the National Institute for Health and Care Research (NIHR) Manchester Biomedical Research Centre (NIHR203308). A.B. is supported by a Wellcome PhD Training Fellowship for Clinicians and the 4Ward North PhD Programme for Health Professionals (223521/Z/21/Z).

This research was made possible through access to data in the National Genomic Research Library, which is managed by Genomics England Limited (a wholly owned company of the Department of Health and Social Care). The National Genomic Research Library holds data provided by patients and collected by the NHS as part of their care and data collected as part of their participation in research. The National Genomic Research Library is funded by the National Institute for Health Research and NHS England. The Wellcome Trust, Cancer Research UK and the Medical Research Council have also funded research infrastructure. This research has been conducted using the UK Biobank Resource under Application Number 90952.

The Australian Undiagnosed Diseases Network (UDN-Aus) acknowledges financial support from the Australian Government’s Medical Research Future Fund (2007567), Australian Genomics and The Centre for Population Genomics (Garvan Institute of Medical Research and Murdoch Children’s Research Institute); funded in part by a Medical Research Future Fund (MRFF) Genomics Health Futures Mission grant (2008820).

## CONFLICTS OF INTEREST

L.M. has received personal consultancy fees from Mendelian Ltd., a rare disease digital health company, outside of the submitted work. No other authors have any conflicts to declare.

## AUTHOR CONTRUBUTIONS

AJ, AB and SB conceptualised the study, analysed data and wrote the manuscript. SB and JL provided supervision. OA, ND, JL, RR, CS and SW analysed data. AJ, SB, SA, EB, AFB, HB, KEC, NC, FE, LE, AF, GG, UK, RM, SM, LM, KM, AM, EP, AP, KP, MR, AR, CLO, RS, SS, HS, FT and EW assisted in clinical data collection. ROK and HT generated data. NW provided technical expertise. All authors read and approved the final version of this manuscript prior to submission.

## DATA AVAILABILITY

Genomic and phenotypic data are available for the 100KGP and individuals who have had WGS through the Genomic Medicine Service in the NGRL. Access to the NGRL may be granted following application via https://www.genomicsengland.co.uk/research/academic/join-research-network, which gives access to the secure GERE. Genomic data used pertain to participants in 100KGP in the Main Programme v.18 and the GMS data v.4. SolveRD data are accessible by application through the RD-CONNECT platform. All data presented in this paper, pertaining to 100kGP participants, were requested for the Airlock transfer through GERE. The paper was submitted for approval by the Genomics England Publication Committee on 25th August 2025 and was approved on 27^th^ August 2025. Access to the Australian Centre for Population Genomics dataset can be requested through contact with the authors. The GRCh38 human genome reference assembly can be accessed at https://www.ncbi.nlm.nih.gov/datasets/genome/GCF_000001405.26/. The GENCODE v.32 comprehensive annotations were accessed within the GERE but can be downloaded from https://www.gencodegenes.org/human/release_32.html. The gnomADv4 genotype VCF files were accessed within the GERE but can also be downloaded from https://gnomad.broadinstitute.org/.

## Notes

### Funding Statement

A.J. and S.B. acknowledge the support of SolveRD. The SolveRD project has received funding from the European Unions Horizon 2020 research and innovation program under grant agreement 779257. This study has been delivered through the National Institute for Health and Care Research (NIHR) Manchester Biomedical Research Centre (NIHR203308). A.B. is supported by a Wellcome PhD Training Fellowship for Clinicians and the 4Ward North PhD Programme for Health Professionals (223521/Z/21/Z). This research was made possible through access to data in the National Genomic Research Library, which is managed by Genomics England Limited (a wholly owned company of the Department of Health and Social Care). The National Genomic Research Library holds data provided by patients and collected by the NHS as part of their care and data collected as part of their participation in research. The National Genomic Research Library is funded by the National Institute for Health Research and NHS England. The Wellcome Trust, Cancer Research UK and the Medical Research Council have also funded research infrastructure. This research has been conducted using the UK Biobank Resource under Application Number 90952. The Australian Undiagnosed Diseases Network (UDN-Aus) acknowledges financial support from the Australian Governments Medical Research Future Fund (2007567), Australian Genomics and The Centre for Population Genomics (Garvan Institute of Medical Research and Murdoch Childrens Research Institute); funded in part by a Medical Research Future Fund (MRFF) Genomics Health Futures Mission grant (2008820).

### Author Declarations

This research was performed under the ethical approvals given by the South Manchester National Health Service (NHS) Research Ethics Committee (REC; 11/H1003/3/AM02

### Summary of Updates

Revision to correct figure order

