## Supplementary Note for "Biallelic variants in *RNU2-2* cause a remarkably frequent developmental epileptic encephalopathy"

### Supplementary Case Reports

REDACTED FOR MEDRXIV

### Specific variants and variant combinations

We have previously shown that heterozygous variation in the 5’ end of RNU2-2 (n.1 to n.67) is highly constrained in gnomAD v4.1^4^. This region contains nucleotides which carry extensive post-transcriptional modifications and are involved in inter-molecular base pairing with U6 and the splicing branchpoint.

Distinct from the dominant *RNU2-2* syndrome, where variants are recurrent, we observed a diverse set of variant combinations in affected cases in this study. All of these variants are rare in gnomAD, however, we do observe some recurrent variants in unrelated families.

Four variants were observed in both the homozygous and compound heterozygous states (n.20G>A, n.28C>G, n.104T>C and n.127G>C)*.* Variant positions in unsolved NDD cases were distributed throughout the primary sequence of *RNU2-2*, apart from the unstructured sequence between stem loop IIb and the Sm binding site (n.85 to n.96) which was devoid of variants in these individuals (Fisher’s exact P=0.0091). Variants in unsolved NDD cases overlapped nucleotides important for snRNA-snRNA interactions between U2 and U6, snRNA-protein interactions within the Sm protein binding site, and U2 secondary loop structures including stem loops IIa, IIb, III and IV.

Recurrent variants in unsolved NDD cases were n.45C>T and n.104T>C, which were each observed in two and three unrelated families, respectively*.* Different substitutions were observed at these nucleotides also (n.45C>G and n.104T>G) in the cohort. The recurrence of variants in affected individuals, and the occurrence of variants at positions which base-pair in the snRNA secondary structure, provide supporting evidence of pathogenicity. For example, three individuals from two unrelated families (Families 4 and 5), all of Pakistani descent, carried the same n.20G>A variant in homozygous state. Haplotype analysis revealed a shared region of homozygosity of 30Mb between these unrelated families, suggesting a common ancestor. This variant was also observed in another individual, in the compound heterozygous state, *in trans* with n.61C>T (Family 12)*.* n.20 base-pairs with n.13 in the U2-2 secondary structure; we observe n.13C>A in one individual (Family 9) in the compound heterozygous state, in trans with n.100T>G, within the Sm site. Interestingly, we observed a recurrent indel in both the homozygous and compound heterozygous state in three unrelated individuals (n.116_127del in families 7 and 40, and n.116_127dup in family 23). We observed four additional indels in unsolved NDD cases. All were in the compound heterozygous state, and all were located in the 3’ end of *RNU2-2*, downstream of n.115. Two of these (n.170_*5del and n.186_*27del) overlapped the 3’ extension of U2-2 pre-snRNA which is known to be transcribed and cleaved post-transcriptionally, and which contains nucleotides important for regulating transcription termination and stability during nuclear export^5^.

Interestingly, we find two variants in the compound heterozygous state (n.3C>T and n.5C>T) which overlap sites at which specific SNVs (n.3C>A and n.5C>A) we had previously linked to the dominant RNU2-2 syndrome^4^. In both cases, the variants in the dominant condition are observed in individuals with milder presentations, have been seen to be transmitted or parental samples were not available to determine de novo origin. In the family carrying the n.5C>T variants, this was inherited by two affected half-siblings from their unaffected mother. Each half-sibling had, in fact, inherited a different variant from each of their unrelated fathers suggesting that the gene carrier rate of this condition may be high.

Variant interpretation is, of course, challenging in a non-coding gene. For individuals with a low U2:U1 transcript ratio, we find evidence for pathogenicity for the following variant combinations: n.41T>A | n.127G>C, n.20G>A | n.61C>T, n.102T>C homozygous, [n.162dup;n.189T>C] | [n.43T>G;n.62T>G], n.183G>C homozygous, n.171_187del | n.186_*27del, n.181G>C homozygous and n.113G>A | n.116_127dup (**Supplementary Table 10)**
