## Extended Data Figure for "Biallelic variants in *RNU2-2* cause a remarkably frequent developmental epileptic encephalopathy"

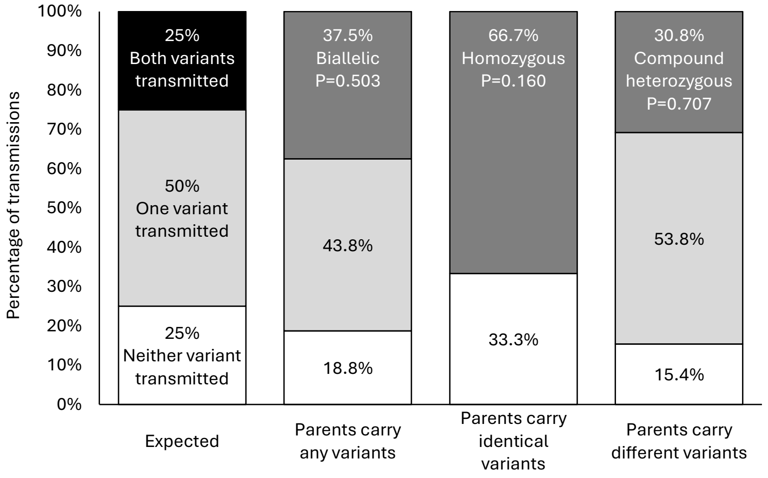


| | Expected | Parents carry any variants | Parents carry identical variants | Parents carry different variants |
|---|---|---|---|---|
| Both variants transmitted | 25 | 6 | 2 | 4 |
| One variant transmitted | 50 | 7 | 0 | 7 |
| Neither variant transmitted | 25 | 3 | 1 | 2 |

**Extended Data Figure 1**| Stacked bar plot showing transmission of heterozygous *RNU2-2* variants from parents to offspring among 100kGP controls.

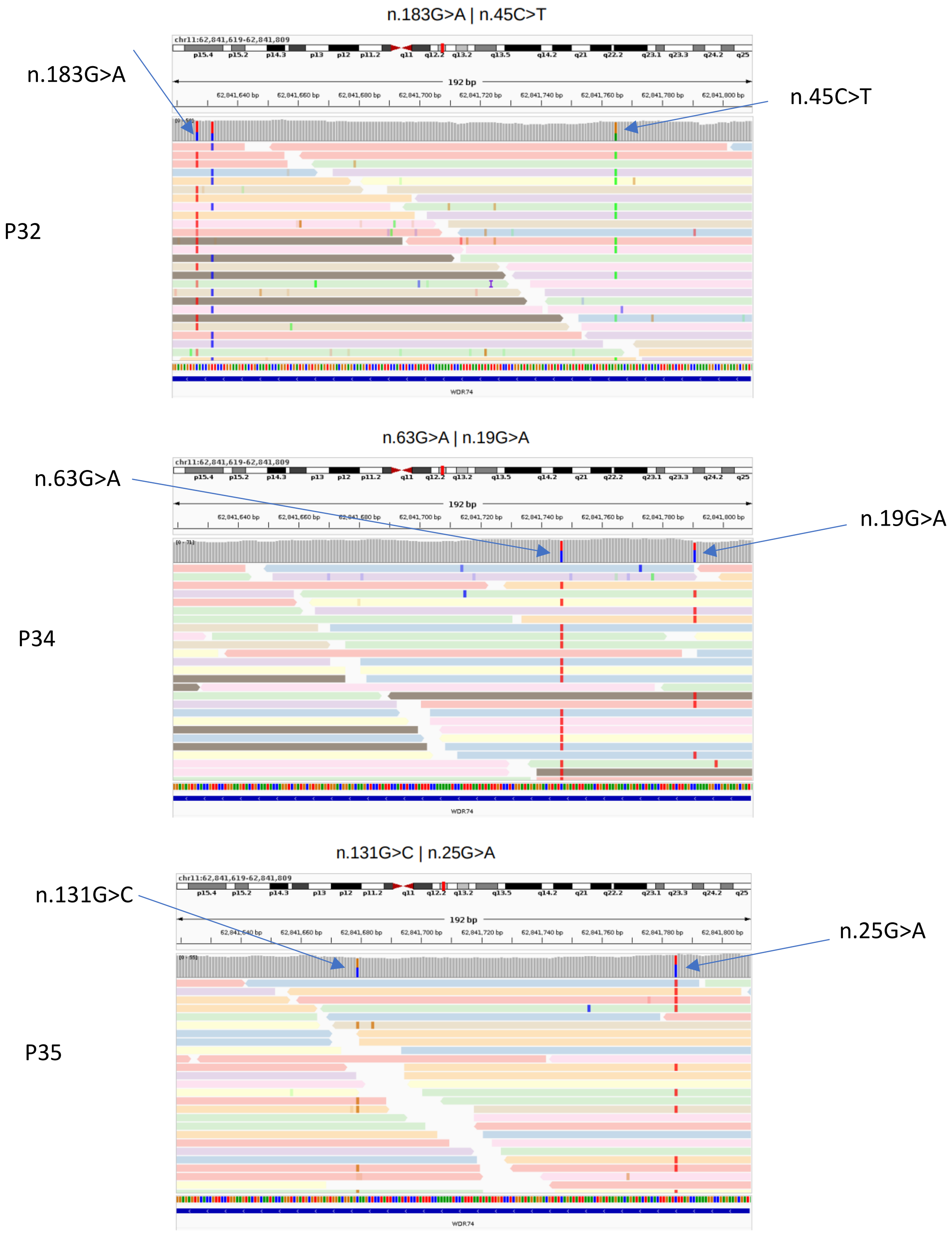


**Extended Data Figure 2**| IGV screenshots of sequencing reads covering *RNU2-2* for individuals with unsolved NDD. Screenshots for three individuals with “phase switch” errors (false negative for compound heterozygosity) in aggV2 are shown. In the top panel, due to the large distance between variants, there is only one informative read (the lowest read in the IGV plot).

RNU2-2 remap chr11 GRCh38

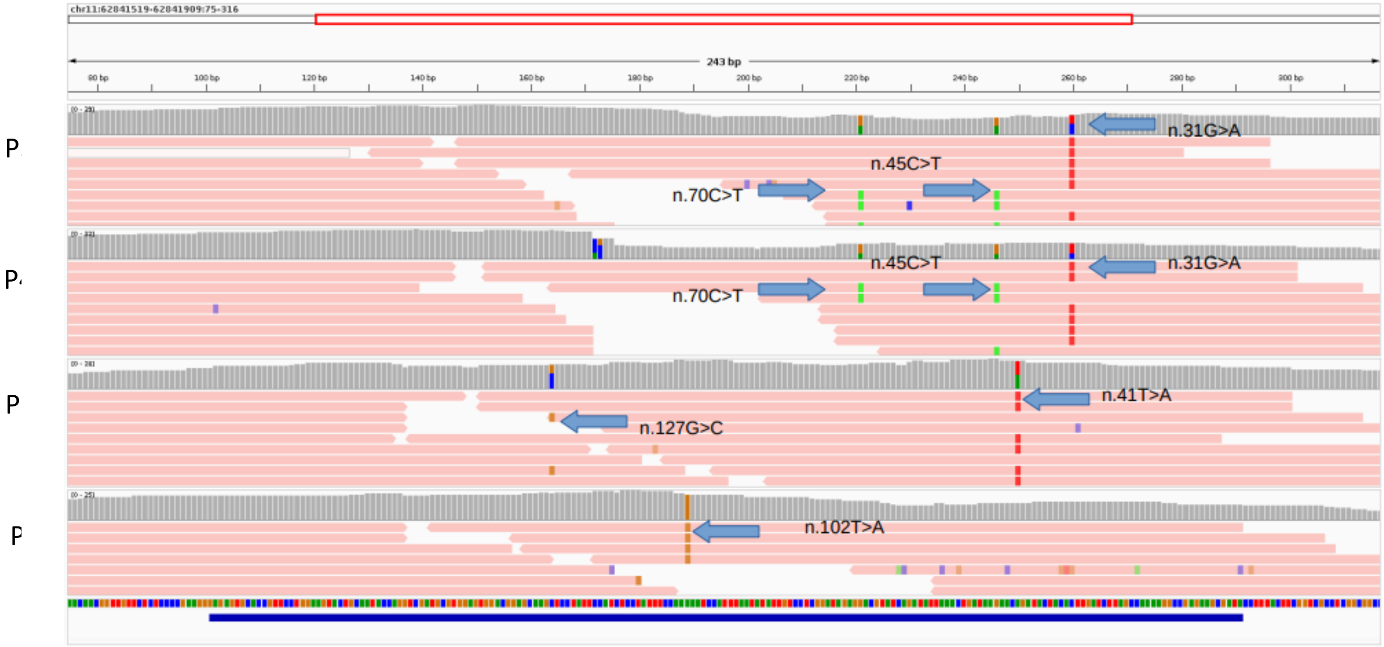


**Extended Data Figure 3**| IGV screenshots of sequencing reads covering *RNU2-2* for individuals with unsolved NDD who had their data aligned to GRCh37, where *RNU2-1* locus is not mapped. The IGV screenshot shows all reads re-mapped to *RNU2-2* in GRCh38 indicating confident variant calls. For P39 and P40, the rare (n.45C>T and n.31G>A) variants are shown whilst a common variant (n.70C>T) is in linkage with n.45C>T.

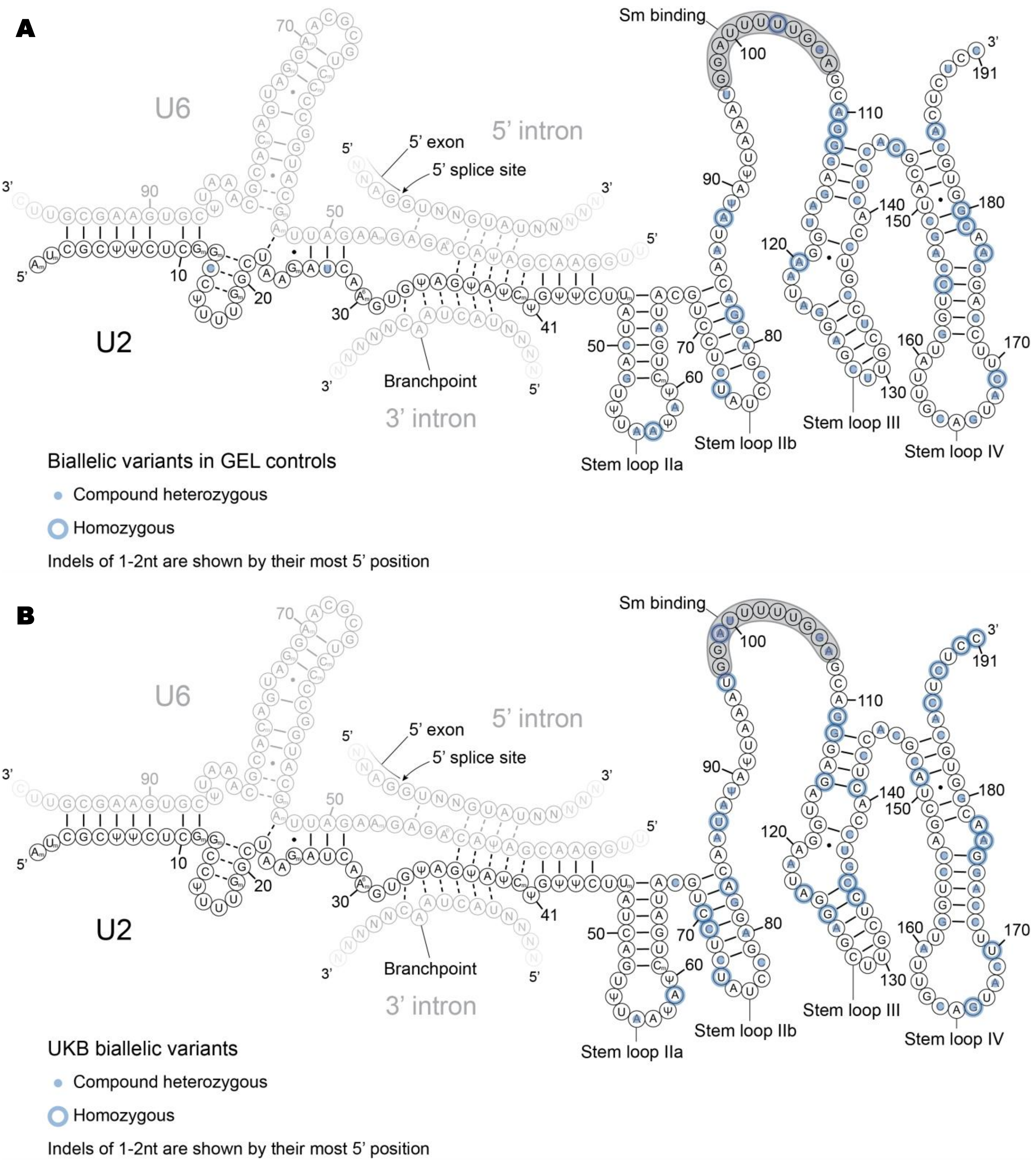


**Extended Data Figure 4**| Secondary structure of U2-2 in complex with U6, with biallelic variants in control cohorts are overlaid. **A)** Biallelic variants in GEL controls. **B)** Biallelic variants in UK Biobank. Secondary structure annotations are shown as in Figure 2B.

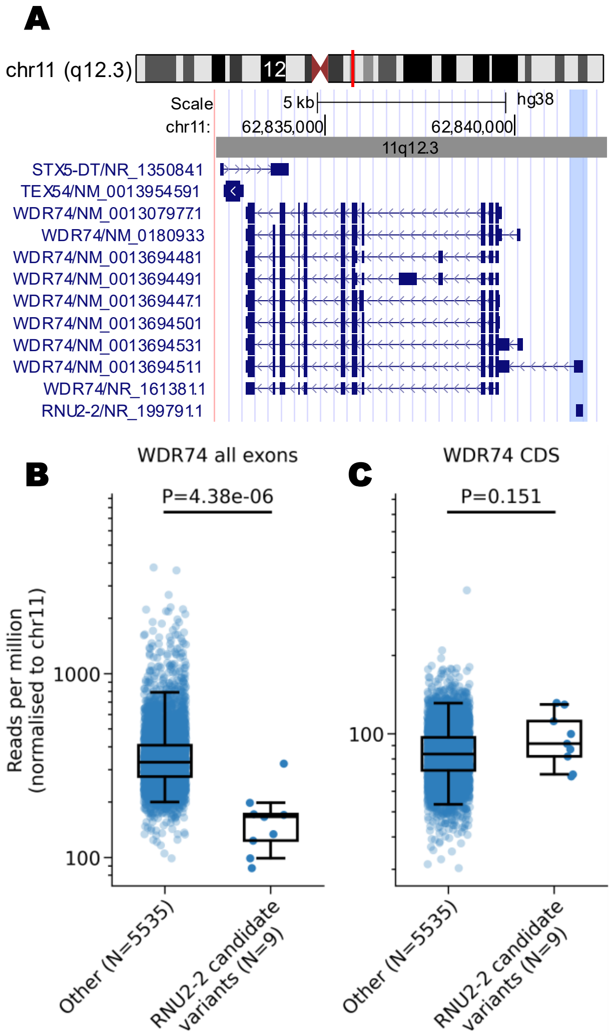


**Extended Data Figure 5**| WDR74 expression in carriers of candidate biallelic variants in *RNU2-2*. **A)** Screengrab from the UCSC genome browser showing the *WDR74* locus and adjacent genomic context (GRCh38 chr11:62,832,122-62,842,440). The blue highlight shows the overlap of *RNU2-2* with the 5' UTR of a single transcript in *WDR74* (NM_001369451.1) **B)** Relative expression of *WDR74* in individuals with candidate biallelic variants in *RNU2-2* versus controls. Reads mapping to all exons in *WDR74* are shown. **C)** As in **B)**, but for reads mapping to CDS exons in *WDR74*. For **B)** and **C)**, read counts are normalised to chromosome 11. P values from two-tailed Mann-Whitney U tests are shown. Box and whisker plots show the median, quartiles, and +/- 1.5 times the interquartile range of the data.

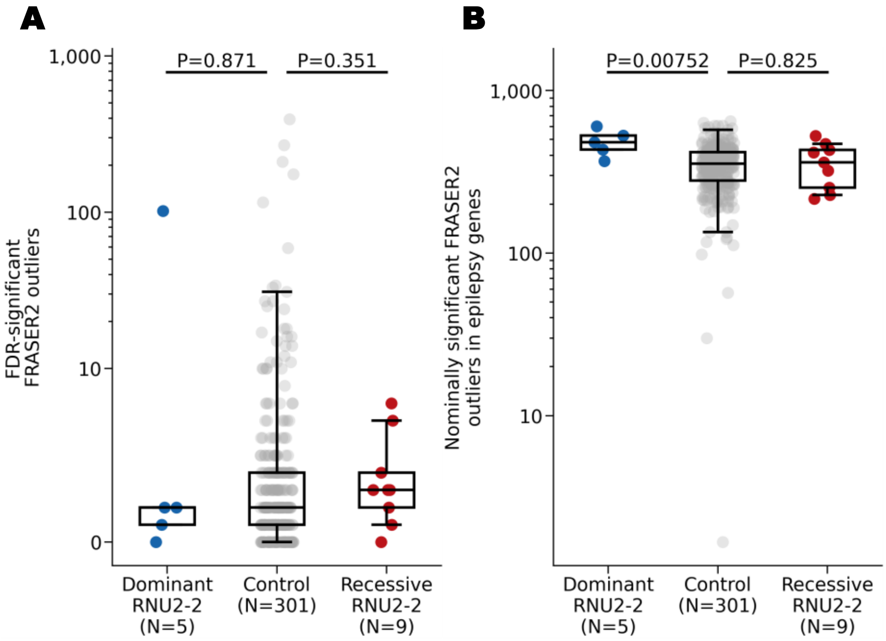


**Extended Data Figure 6**| Aberrant splicing events from RNA sequencing data in individuals with candidate variants in *RNU2-2* versus controls. **A)** FDR-significant (FDR P<0.10) FRASER2 splicing outliers. **B)** Nominally significant (unadjusted P<0.05) FRASER2 outliers in known monogenic epilepsy genes from PanelApp Australia. In each panel, data are shown for individuals with heterozygous variants pathogenic for the dominant *RNU2-2* disorder (blue circles) (N=5), non-NDD controls (grey circles) (N=301), and individuals with unsolved NDD and candidate biallelic variants in *RNU2-2* (red circles) (N=9). P values from two-tailed Mann-Whitney U tests are shown. Box and whisker plots show the median, quartiles, and +/- 1.5 times the interquartile range of the data.

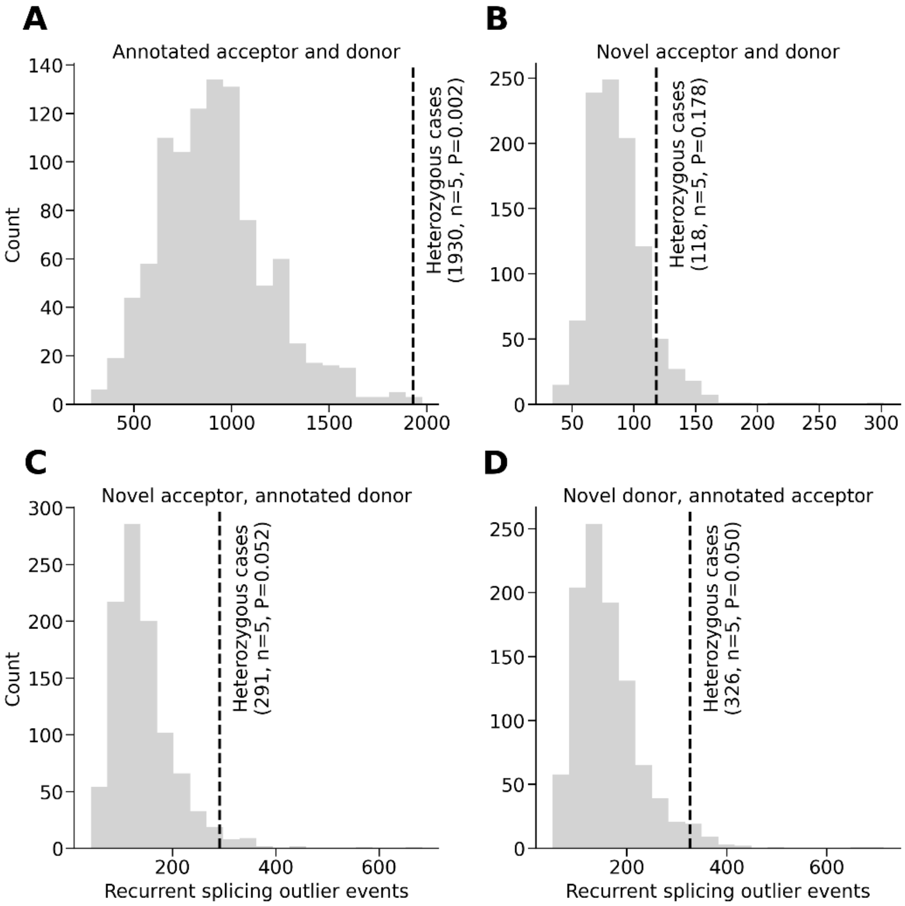


**Extended Data Figure 7**| Recurrent splicing events in individuals with heterozygous variants pathogenic for the dominant *RNU2-2* disorder. **A)** Aberrant splicing events at annotated splice junctions. **B)** Splice junctions containing novel splice acceptor sites and annotated splice donor sites. **C)** Novel splice donor and acceptor sites **D)** Novel splice donor sites and annotated splice acceptor sites. Histograms show the number of identical nominally significant FRASER2 aberrant splicing events observed in >1 individual from 1000 random samples of 5 controls. The dashed black lines show the number of identical nominally significant FRASER2 aberrant splicing events observed in >1 individual among 5 cases with pathogenic heterozygous variants in *RNU2-2*. The number of recurrent splicing events is given in parentheses. Two-sided bootstrap P values are shown.

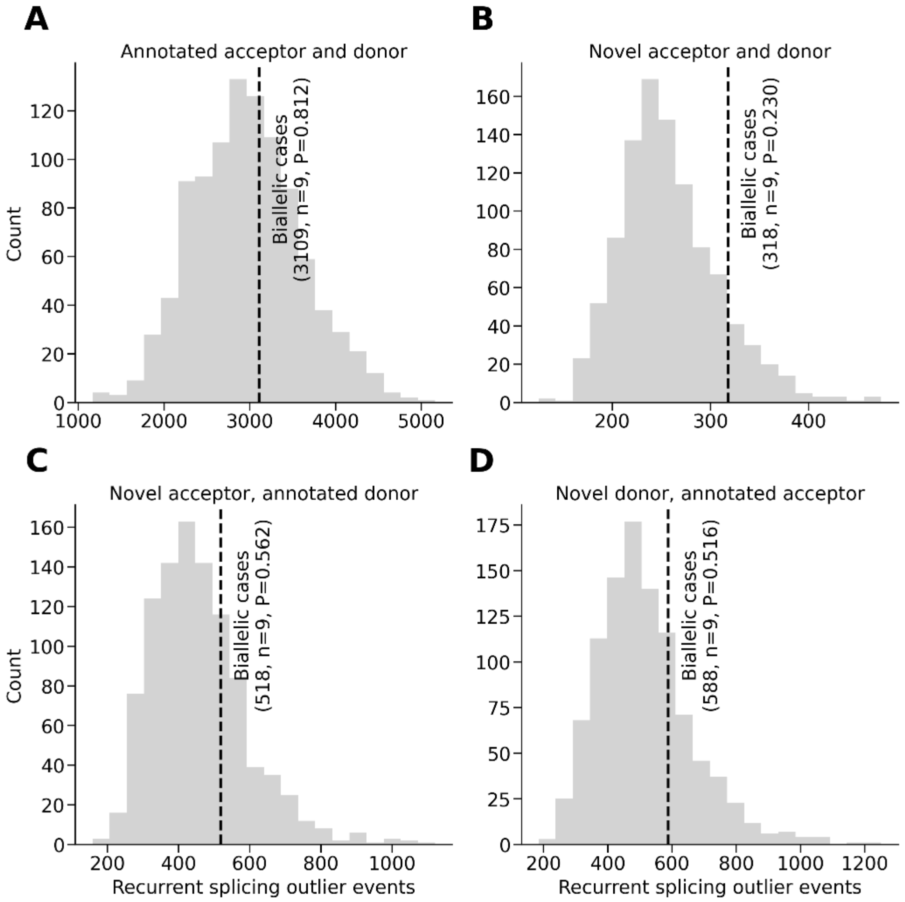


**Extended Data Figure 8**| Recurrent splicing events in individuals with candidate biallelic variants in *RNU2-2*. Panels **A)**, **B)**, **C)** and **D)** are arranged as in Extended Data Figure 7. Histograms show the number of identical nominally significant FRASER2 aberrant splicing events observed in >1 individual from 1000 random samples of 9 controls. The dashed black lines show the number of identical nominally significant FRASER2 aberrant splicing events observed in >1 individual among 9 cases with candidate biallelic variants in *RNU2-2*. The number of recurrent splicing events is given in parentheses. Two-sided bootstrap P values are shown.

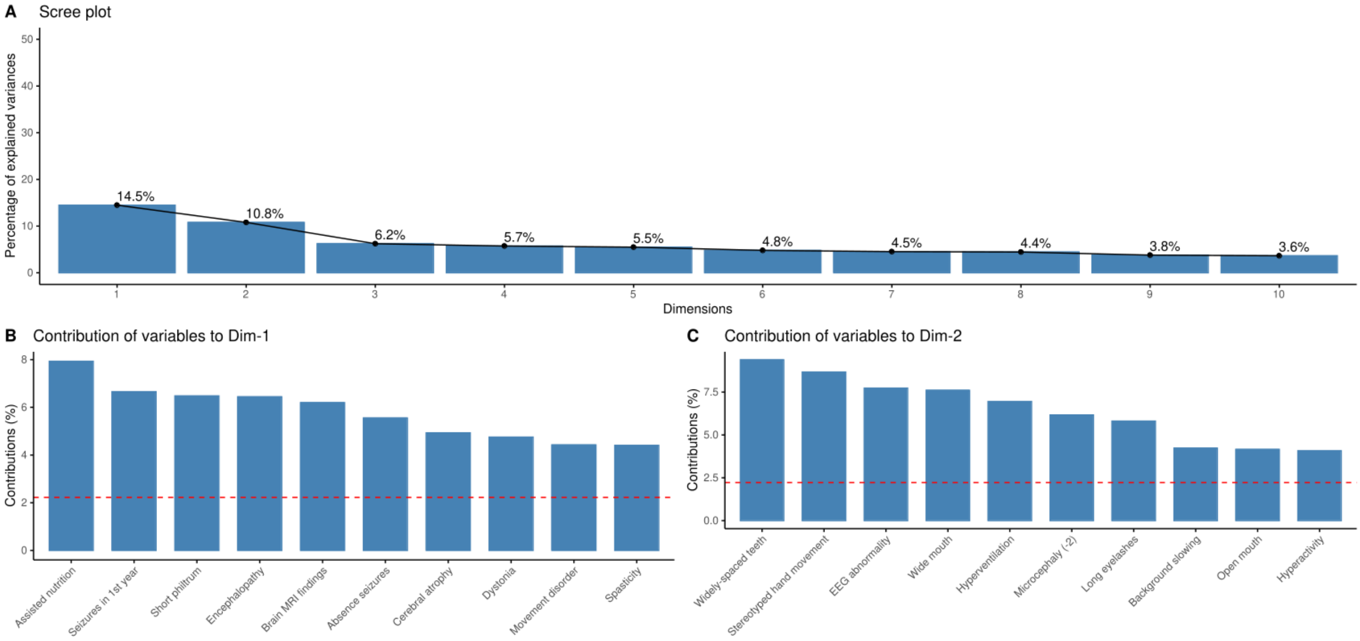


**Extended Data Figure 9**| Details of the PCA analysis comparing HPO term frequency in the recessive and dominant *RNU2-2* disorders. **A)** Proportion of the variance explained by the first ten principal components. **B)** The relative contribution of the 10 most influential HPO terms to the first principal component. **C)** The relative contribution of the 10 most influential HPO terms to the second principal component. The red dashed line indicates the expected value if each contribution was uniform.

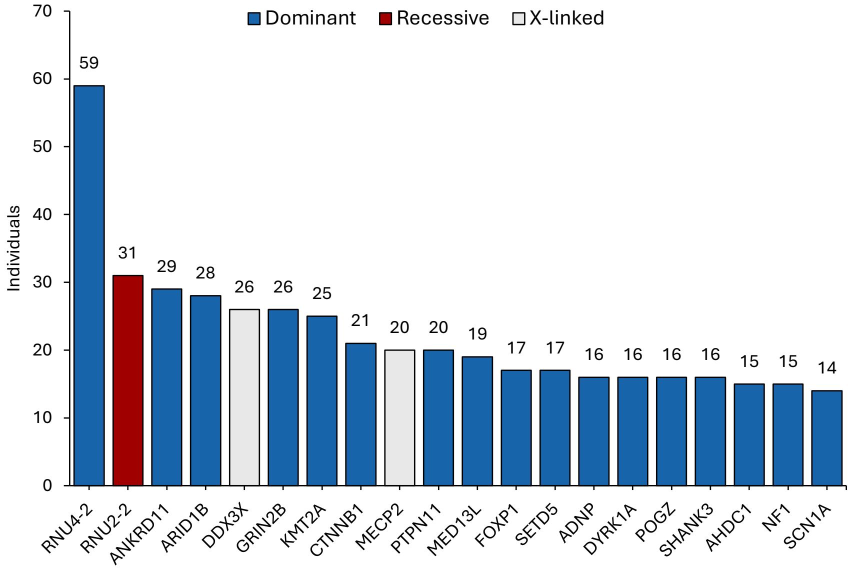


**Extended Data Figure 10**| Bar plot showing the 20 most frequent diagnoses on the R29 Intellectual Disability panel for individuals in the 100kGP recruited with NDD (N=10,987). Counts for the *RNU2-2* and *RNU4-2* disorders were based on genotyping (see Methods). Counts for all other genes were taken from exit questionnaire data. Genes causal for dominant disorders are shown in blue, for X-linked disorders in grey, and for recessive disorders in red.
